# Blood-based epigenome-wide analyses of chronic low-grade inflammation across diverse population cohorts

**DOI:** 10.1101/2023.11.02.23298000

**Authors:** Robert F. Hillary, Hong Kiat Ng, Daniel L. McCartney, Hannah R. Elliott, Rosie M. Walker, Archie Campbell, Felicia Huang, Kenan Direk, Paul Welsh, Naveed Sattar, Janie Corley, Caroline Hayward, Andrew M. McIntosh, Cathie Sudlow, Kathryn L. Evans, Simon R. Cox, John C. Chambers, Marie Loh, Caroline L. Relton, Riccardo E. Marioni, Paul D. Yousefi, Matthew Suderman

**Affiliations:** Centre for Genomic and Experimental Medicine, Institute of Genetics and Cancer, University of Edinburgh, Edinburgh, UK; Lee Kong Chian School of Medicine, Nanyang Technological University, Clinical Sciences Building, Singapore, Singapore; MRC Integrative Epidemiology Unit at the University of Bristol, Bristol, United Kingdom; Population Health Sciences, Bristol Medical School, University of Bristol, Bristol, United Kingdom; School of Psychology, University of Exeter, Exeter, United Kingdom; MRC Unit for Lifelong Health and Ageing, University College London, London, United Kingdom; Imperial Clinical Trials Unit, School of Public Health, Imperial College London, London, United Kingdom; School of Cardiovascular and Metabolic Health, BHF Glasgow Cardiovascular Research Centre, University of Glasgow, Glasgow, United Kingdom; Lothian Birth Cohort studies, Department of Psychology, University of Edinburgh, Edinburgh, United Kingdom; Medical Research Council Human Genetics Unit, Institute of Genetics and Cancer, University of Edinburgh, Edinburgh, United Kingdom; Division of Psychiatry, University of Edinburgh, Royal Edinburgh Hospital, Edinburgh, United Kingdom; Centre for Clinical Brain Sciences, Edinburgh Imaging and UK Dementia Research Institute, University of Edinburgh, Edinburgh, United Kingdom; British Heart Foundation Data Science Centre, Health Data Research UK, London, United Kingdom; Health Data Research UK, London, United Kingdom; Department of Epidemiology and Biostatistics, School of Public Health, Imperial College London, St Mary’s Campus, London, United Kingdom; National Skin Centre, Singapore, Singapore; Genome Institute of Singapore, Agency for Science, Technology and Research, Singapore, Singapore

**Author notes:** Correspondence: Riccardo E. Marioni, Paul D. Yousefi or Matthew Suderman.

## Abstract

Chronic inflammation is a hallmark of ageing and age-related disease states. The effectiveness of inflammatory proteins such as C-reactive protein (CRP) in assessing long-term inflammation is hindered by their phasic nature. DNA methylation (DNAm) signatures of CRP may act as more reliable markers of chronic inflammation. We show that inter-individual differences in DNAm capture 50% of the variance in circulating CRP (N=17,936, Generation Scotland). We develop a series of DNAm predictors of CRP using state-of-the-art algorithms. An elastic net regression-based predictor outperformed competing methods and explained 18% of phenotypic variance in the LBC1936 cohort, doubling that of existing DNAm predictors. DNAm predictors performed comparably in four additional test cohorts (ALSPAC, HELIOS, SABRE, LBC1921), including individuals of diverse genetic ancestry and from different age groups. The newly-described predictor surpassed assay-measured CRP and a genetic score in its associations with 26 health outcomes. Our findings forge new avenues for assessing chronic low-grade inflammation in diverse populations.

## 1 Introduction

Chronic low-grade inflammation is a common feature of many age-related disease states, including heart disease, stroke and type 2 diabetes^1,2^. C-reactive protein (CRP) is a sensitive marker of systemic inflammation^3^. However, individual measurements can fluctuate substantially following infection or injury. Therefore, single time-point measures of CRP in clinical settings may provide an incomplete index of an individual’s long-term inflammatory status^4^. Identifying inflammatory biomarkers with enhanced temporal stability could improve patient stratification and facilitate robust health outcome testing.

DNA methylation (DNAm) is a reversible epigenetic mechanism in which methyl groups bind to DNA, most commonly within cytosine-guanine dinucleotides (CpG sites). DNAm is affected by a confluence of genetic and environmental factors and is known to regulate gene expression levels^5^. Two large-scale epigenome-wide association studies (EWAS) have observed >1,000 CpG sites across the genome that associate with blood CRP levels^6,7^. DNAm predictors of CRP levels have been constructed using weighted linear combinations of these CpG sites and explain approximately 10% of inter-individual variation in circulating CRP^7,8^. They show greater longitudinal stability than measured CRP as well as stronger associations with cognitive and cardiometabolic health outcomes^6,8–10^, indicating they are likely to be robust to short-term CRP fluctuations.

Addressing the following points will be critical to further extending our understanding of how DNAm informs the biology and prediction of CRP. There is a need to estimate the expected proportion of variance in CRP captured by genome-wide DNAm probes. Variance component estimates would guide an upper bound for the amount of variance in CRP that can be captured by DNAm predictors, as well as inform the molecular architecture of CRP regulation. Existing predictors are constructed using weights from individual linear regressions. The approach neglects correlations between CpG sites and unknown confounding influences. Bayesian regression methods and several commonly-employed feature selection methods, such as penalised regression and principal component analysis (PCA), may overcome these limitations^11,12^. Furthermore, studies examining the relationship between DNAm and CRP are primarily restricted to adults of European ancestry. It is unclear if DNAm predictors of CRP are generalisable to other stages of the life-course (e.g. childhood and later-life) or across genetically diverse individuals.

We address four primary objectives by leveraging blood-based methylation and CRP measurements across six diverse cohorts (N_range_=170 to 17,936). First, we bolster biological insights into the relationship between DNAm and CRP by conducting a large-scale EWAS of CRP in the family-based study Generation Scotland (GS, N=17,936). We also employ two complementary methods, restricted maximum likelihood estimation and Bayesian penalised regression, to estimate the proportion of inter-individual variation in CRP attributable to genome-wide DNAm^11,13^. Second, we address unmet prediction efforts by applying three common feature selection and transformation methods to develop new DNAm predictors of CRP levels. They are elastic net regression, Bayesian penalised regression and principal component analysis. Third, we compare their predictive performances against one another and to existing predictors in the literature. GS serves as the training cohort in prediction analyses. Five diverse test cohorts are employed: Avon Longitudinal Study of Parents and Children (ALSPAC, mother and child pairs), Health for Life in Singapore (HELIOS, adults of self-reported Chinese, Malay or Indian ethnicity), Southall and Brent REvisited (SABRE, adult males of self-reported European or South Asian ethnicity) and the Lothian Birth Cohorts of 1921 and 1936 (community-dwelling older adults). Fourth, we compare newly-described DNAm predictors of CRP against assay-measured CRP in their associations with 26 cardiometabolic risk factors and health outcomes. **Fig. 1** shows a visual summary of the study design.

**Fig. 1.**
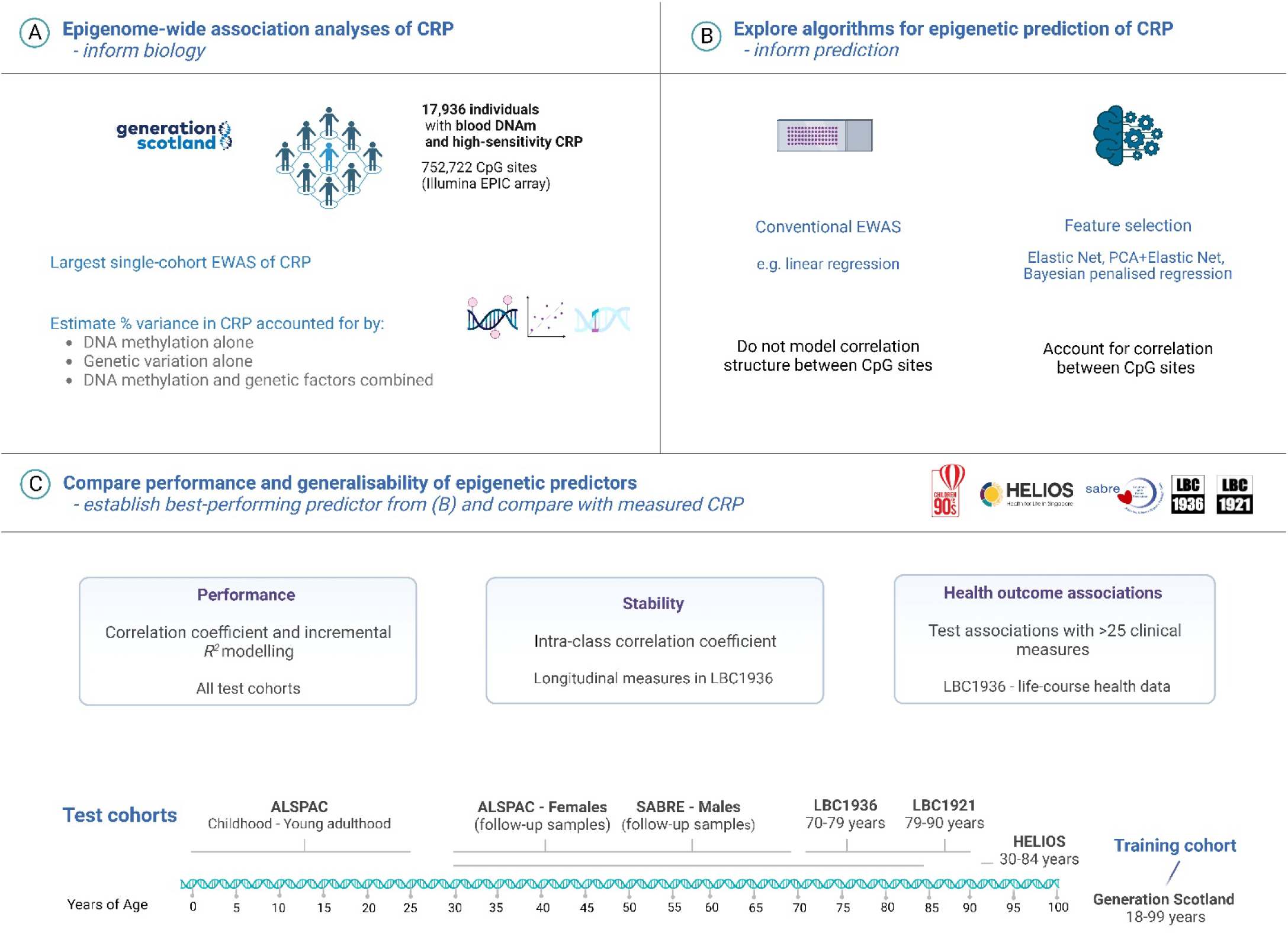
Blood epigenome-wide analyses on C-reactive protein levels across a diverse set of population cohorts. (A) There were 17,936 individuals in Generation Scotland with complete high-sensitivity CRP measurements and genome-wide DNAm profiling. This allowed for a large epigenome-wide scan for associations between differential DNA methylation and blood CRP levels, alongside a variance component analysis of molecular phenotypes and CRP. (B) A suite of feature selection and transformation methods were implemented to develop new DNAm predictors of CRP. These methods account for the correlation structure between features (CpG sites) and may offer improved predictive performances over existing methods (i.e. methylation risk scores with weights from linear regression models). (C) The predictive performances of CRP predictors derived from feature selection methods in (B) were compared against existing predictors. The five test cohorts harboured cross-sectional samples that encompass the life-course (i.e. cord blood samples and childhood through to later-life), adult males and females, and individuals from different ethnic backgrounds and countries of residency. Of the test cohorts, the Lothian Birth Cohort 1936 was selected for health outcome testing given that the study population was at elevated risk for age-related disease states when compared to other cohorts and sub-groups. It also constituted a larger analytical sample than the Lothian Birth Cohort 1921. ALSPAC, Avon Longitudinal Study of Parents and Children; CpG, cytosine-phosphate-guanine dinucleotide; CRP, C-reactive protein; DNAm, DNA methylation; EWAS, epigenome-wide association study; GS, Generation Scotland; HELIOS, Health for Life in Singapore; LBC1921, Lothian Birth Cohort 1921; LBC1936, Lothian Birth Cohort 1936; SABRE, Southall And Brent REvisited. Image created with Biorender.com.

## 2 Methods

### 2.1 Cohort studies

DNA methylation preparation and CRP quantification across all cohorts are detailed in full in **Additional file 1**. To align with the previous literature, CRP levels were log-transformed after adding a constant of 0.01 to prevent undefined values. Measurements that were outside the median value ±4 times the standard deviation were excluded^6^.

#### 2.1.1 Training Cohort: Generation Scotland

Generation Scotland: Scottish Family Health Study (GS) is a family-structured, population-based cohort study of >24,000 Scottish individuals^14,15^. Recruitment took place between 2006 and 2011. Blood draws were taken during a clinical visit at the study baseline alongside detailed health, lifestyle, cognitive and sociodemographic data. Whole-blood DNAm was measured using the Illumina Infinium MethylationEPIC array. Serum CRP levels (mg/L) were quantified at the University of Glasgow using a commercial high-sensitivity assay on an automated analyser (c311, Roche Diagnostics, UK). There were 17,936 individuals with genome-wide DNAm and CRP measurements following quality control and 752,722 CpG sites were available for analyses (**Additional file 1**).

#### 2.1.2 Test Cohorts

Here, we will present a brief description of each test cohort including the number of individuals with paired CRP and DNAm measurements. Complete information on the test cohorts is available in **Additional file 1**. **Table 1** shows data on demographics and CRP measurements for each test cohort.

**Table 1.** Summary of demographics and C-reactive protein measurement across six diverse cohorts.

| *Sample* | *N* | *Age (years)*<br>*Mean, SD* | *Sex*<br>*N Female, %*<br>*Female* | *CRP (mg/L)*<br>*Mean, SD* | *Assay* |
| --- | --- | --- | --- | --- | --- |
| ***ALSPAC*** |  |  |  |  |  |
| Age 0 | 389 | - | 198, 50.1% | 0.2, 0.9 | High-sensitivity |
| Age 9 | 336 | 9.8, 0.3 | 163, 48.5% | 0.6, 1.1 | High-sensitivity |
| Age 15 or 17 | 945 | 17.1, 1.1 | 492, 52.1% | 1.1, 1.8 | High-sensitivity |
| Age 24 | 745 | 24.5, 0.8 | 365, 49.0% | 1.7, 2.5 | High-sensitivity |
| Mothers (follow-up) | 773 | 47.8, 4.3 | 773, 100% | 1.8, 2.2 | High-sensitivity |
| ***Generation Scotland*** |  |  |  |  |  |
| Whole sample | 17936 | 47.5, 14.9 | 10536, 58.7% | 3.3, 2.9 | High-sensitivity |
| ***HELIOS*** |  |  |  |  |  |
| Chinese Ethnicity | 1778 | 54.6, 11.8 | 729, 61.9% | 1.5, 3.8 | Wide-range |
| Malay Ethnicity | 242 | 55.4, 11.1 | 148, 61.2% | 2.9, 4.9 | Wide-range |
| Indian Ethnicity | 225 | 51.3, 11.8 | 126, 56.0% | 4.4, 7.6 | Wide-range |
| ***SABRE*** |  |  |  |  |  |
| European ethnicity | 315 | 69.9, 6.3 | 0, 0% | 3.6, 7.9 | High-sensitivity |
| South Asian ethnicity | 273 | 69.3, 6.2 | 0, 0% | 2.7, 3.8 | High-sensitivity |
| ***LBC1936*** |  |  |  |  |  |
| Age 70 (W1) | 885 | 69.6, 0.8 | 439, 49.6% | 5.3, 6.9 | Low-sensitivity |
| Age 73 (W2) | 756 | 72.5, 0.7 | 361, 47.8% | 3.0, 5.5 | High-sensitivity |
| Age 76 (W3) | 536 | 76.3, 0.7 | 258, 48.1% | 3.1, 4.8 | High-sensitivity |
| Age 79 (W4) | 492 | 79.3, 0.6 | 240, 48.8% | 2.5, 5.6 | High-sensitivity |
| ***LBC1921*** |  |  |  |  |  |
| Age 87 (W3) | 170 | 86.6, 0.4 | 92, 54.1% | 5.5, 7.2 | Low-sensitivity |
| Age 90 (W4) | 49 | 90.2, 0.1 | 29, 59.2% | 8.4, 11.7 | Low-sensitivity |

Low-sensitivity assays could not reliably detect values below 3 mg/L. These values were set to 1.5 mg/L in line with previous LBC publications<sup>8</sup> and may have contributed to altered distribution properties at time-points and cohorts that used low-sensitivity platforms. ALSPAC, Avon Longitudinal Study of Parents and Children; CRP, C-reactive protein; HELIOS, Health for Life in Singapore; LBC, Lothian Birth Cohorts; SABRE, Southall and Brent REvisited; SD, standard deviation; W, Wave.

##### Avon Longitudinal Study of Children and Parents

Pregnant women who were residing in Avon, UK and with expected dates of delivery from April 1, 1991 to December 31, 1992 were invited to take part in The Avon Longitudinal Study of Children and Parents (ALSPAC)^16–18^. The analytical sample in our study included 773 mothers who had DNAm and CRP measured 18 years after the baseline (i.e. after the study pregnancy)^19^. Four longitudinal measurements of DNAm and CRP were available for the children at the following ages (in years): age 0 (cord blood), 9, 15 or 17, and 24. The number of samples available at each time-point was 389, 336, 945 and 745, respectively. There were 483,068 CpG sites available for testing (**Additional file 1**).

##### Health for Life in Singapore

HELIOS is a population-based cohort of approximately 10,000 Asian men and women living in Singapore. The cohort comprises Singapore citizens or Permanent Residents aged 30–84 years old and excludes pregnant and breastfeeding women, those with major illness requiring hospitalisation or surgery, those who received cancer treatment in the past year or those who participated in drug trials within the month prior to recruitment. Ethnicity was based on self-report data and agreed closely with genetically-determined ancestry^20^. The three primary self-reported ethnicities are: (i) Chinese and other East Asian (Chinese), (ii) Malay and other South-East-Asian (Malay) and (iii) South Asian (Indian and other countries from Indian subcontinent). There were 1,778, 242 and 225 individuals within the Chinese, Malay and Indian ethnicity groups who had paired DNAm and CRP measurements, respectively. The HELIOS dataset contained 837,722 CpG sites (**Additional file 1**).

##### Southall And Brent REvisited

Southall And Brent REvisited (SABRE) is a population-based study that includes 1,711 first-generation South Asian migrants and 1,762 European-origin individuals, all resident in the UK. Recruitment occurred between 1988 and 1991 in West London, UK. Follow-up clinics were held approximately 20 years later between 2008 and 2013^21,22^. Follow-up samples from male participants were used in the present study. Paired DNAm and CRP measurements were available for 588 male participants at the follow-up visit and comprised 315 and 273 participants of self-reported European and South Asian ethnicity, respectively. The final SABRE DNAm dataset contained 484,781 CpG sites (**Additional file 1**).

##### The Lothian Birth Cohorts of 1921 and 1936

The Lothian Birth Cohorts of 1921 (LBC1921; N=550) and 1936 (LBC1936; N=1,091) are longitudinal studies of healthy aging. There were five waves of data collection for LBC1921 at mean ages of 79, 83, 87, 90 and 92 years. The LBC1936 has completed six waves of testing at mean ages of 70, 73, 76, 79, 82 and 86 years^23,24^. Low-sensitivity CRP measures were present at Wave 1 of LBC1936 and Waves 3 and 4 of LBC1921. High-sensitivity CRP measurements were available at Waves 2, 3 and 4 of LBC1936. The number of individuals contributing to our analyses were 885, 756, 536 and 492 at Waves 1, 2, 3 and 4 of the LBC1936, and 170 and 49 at Waves 3 and 4 of the LBC1921, respectively. There were 459,309 CpG sites following quality control (**Additional file 1**).

### 2.2 Epigenome-wide association studies on CRP levels

#### 2.2.1 Linear regression

Our study analysed primary data from 17,936 individuals in GS. For comparability, we approximated the modelling approach of a recent EWAS meta-analysis of CRP by Wielscher *et al.*, which was performed in 30 independent studies (N=22,774)^6^. Specifically, 752,722 CpGs were entered separately into linear regression models using the *lm* function in base R. A basic model and fully-adjusted model were considered. The basic regression model directly matched the strategy of Wielscher *et al.* as follows:

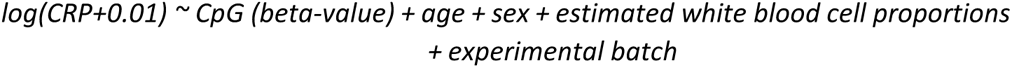

White blood cell proportions were estimated via the Houseman method^25^. The cell types were B cells, CD4^+^ T cells, CD8^+^ T cells, granulocytes, monocytes and natural killer cells. The proportions of granulocytes were omitted from regression models to avoid collinearity given that the proportions of all six cell types sum to 1. Look-up analyses of associations identified in the basic model were performed via the EWAS Catalog^26^.

The fully-adjusted model was unique to our study and further considered population structure and five common lifestyle factors given their possible confounding influences on inflammatory and methylation profiles. The lifestyle factors were alcohol consumption, body mass index, deprivation (Scottish Index of Multiple Deprivation), a methylation-based smoking score (EpiSmokEr)^27^, and years of education, and were selected to align with previous publications in the GS cohort^28,29^. Multidimensional scaling (MDS) was applied to GS genotype data to obtain an estimate of population structure. The first 20 MDS components were fitted as fixed-effect covariates. Demographic and covariate data for GS are presented in **Additional file 2: Table S1**. Their associations with CRP are shown in **Additional file 2: Table S2**. The fully-adjusted model was as follows:

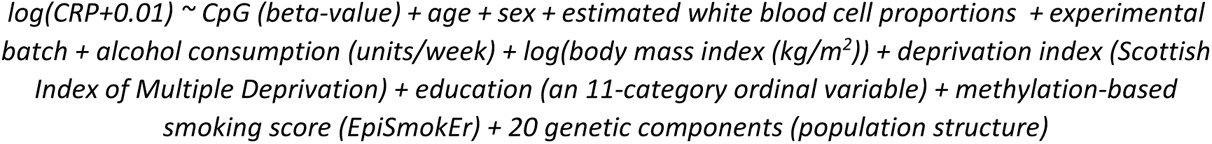

#### 2.2.2 Bayesian penalised regression

Models considering linear associations between outcomes and individual molecular traits do not account for correlation structures within molecular datasets and omitted variable bias^30^. A number of methods have been proposed to overcome these limitations, which include Bayesian penalised regression. The BayesR+ framework implements Bayesian penalised regression and Gaussian mixture-based variance partitioning to inform the molecular architecture of phenotypes^11^. The joint and conditional effects of all 752,722 CpG sites on blood CRP levels were examined. Linear regression models were used to adjust log-transformed CRP levels for age and sex. They were also used to regress CpG beta-values on age, sex, WBC proportions and experimental batch. Residuals from the regressions were scaled to mean zero and unit variance. The prior mixture variances were set to 0.001, 0.01 and 0.1, which corresponds to CpGs that capture 0.1%, 1% and 10% of variation in CRP^6,7^. Details of posterior sampling are provided in **Additional file 3**. Probes with a posterior inclusion probability ≥20% were first identified as lead probes. We then grouped probes that were within 2.5kb of, and highly correlated (absolute Pearson’s correlation coefficient >0.5) with, a lead probe. Groups where the combined posterior inclusion probability was >80% were considered of relevance and the lead probe was highlighted for clarity.

### 2.3 Estimating proportion of variance in CRP levels attributable to genetics and DNAm

BayesR+ was one of two methods implemented in variance component estimation. Here, the same pipeline from EWAS was applied. Variance components estimates were taken as the mean of the sum of squared standardised posterior effect sizes across 1,000 iterations. The 2.5%ile and 97.5%ile (iteration rank 25 and 975) formed the lower and upper bounds of the 95% credible interval, respectively.

Variance partitioning was also performed using OSCA (OmicS-data-based Complex trait Analysis) software^13^. The software estimates the phenotype variance captured by a given set of genome-wide molecular trait or omic data (e.g. DNAm) by first constructing an omics relationship matrix (ORM) using all input probes. The ORM describes the covariance pattern across individuals. A univariate linear mixed model is then implemented and fits the ORM as a random effect component. The variance attributed to the molecular component is obtained via restricted maximum likelihood (REML) estimation. Phenotype and methylation data were adjusted as per the BayesR+ strategy. The ORM was constructed from the residuals of all CpG sites (n=752,722 sites).

A combined genetic-epigenetic model was also performed in BayesR+ and OSCA to enable an estimate of additive and independent effects of DNAm and genetic data over CRP (**Additional file 3**).

### 2.4 DNAm prediction of CRP levels

Three primary methods were used to build weighted linear predictors of CRP from genome-wide DNAm: (1) elastic net regression, (2) Bayesian penalised regression and (3) PCA combined with elastic net regression (‘PCA+elnet’). The training dataset in all instances was GS (N=17,936). An adjusted BayesR+ pipeline was applied in prediction when compared to EWAS and variance partitioning. Phenotype and methylation data were adjusted for potential confounders in EWAS and variance component analyses given that the aim was biological inference. Here, log-transformed CRP and DNAm were not adjusted prior to entry in BayesR+; however, they were scaled to mean zero and unit variance. This enabled us to capture reciprocal and external influences on DNAm and CRP, aiding in external prediction efforts. Further, the number of probes in prediction models was restricted to those present in ALSPAC, GS, LBC and SABRE following quality control (n=374,785 sites). A weighted linear combination of these probes and their effect sizes was used to compute the resultant Bayesian penalised regression-based predictor in all test cohorts (see **Data Availability**). Of note, this probe set (n=374,785) was used to develop our predictors. However, we also wished to understand how missing CpG sites would impact prediction reflecting a real-world scenario where external cohorts utilise our weights but may not contain all of the same CpG sites. Whereas, ALSPAC, GS, LBC and SABRE contained all 374,785 sites (by design), the HELIOS dataset possessed 374,592 of the sites thereby lacking 193 sites in the Bayesian predictor.

Elastic net regression is a commonly-employed technique to develop DNAm predictors of human traits^31^. Regression models were run using the R package *glmnet*^32^. Log-transformed CRP values were entered as the dependent variable and CpG beta-values (scaled to mean zero and unit variance) served as the independent variables (n=374,785 sites). The mixing parameter was set to 0.5 (a common default parameter for elastic net models) and twenty-fold cross-validation was applied. The model with the lambda value that corresponded to the minimum mean cross-validated error was selected. The optimal model contained 1,468 probes (**Additional file 2: Table S3**). HELIOS harboured 1,466 of these sites while all other test cohorts contained all 1,468 sites.

The third method we considered was PCA with elastic net regression. Higgins-Chen *et al*. enhanced the reliability of epigenetic age estimates by combining PCA with elastic net^12^. In this approach, PCA is applied to all CpG sites of interest in a training dataset in order to identify sets of multi-collinear sites. Elastic net regression is then used to identify an optimal combination of PCs that can predict trait values in external test samples. Here, we first truncated the training DNAm dataset to probes that were deemed significant in a CRP EWAS from Wielscher *et al.* The authors identified 1,511 independent sites at a *p*-value threshold of 1 x 10^-7^. There were 1,379 of these 1,511 sites in our training dataset (i.e. GS). All test cohorts also contained the same set of 1,379 sites including HELIOS. We further considered an additional series of *p*-value thresholds (beyond 1 x 10^-7^) for probe filtering as sensitivity analyses (see **Section 3.5**). PCA was performed using the *prcomp* function in R and training data were mean-centred but not scaled as described by Higgins-Chen *et al*^12^. Elastic net regression was implemented using the parameters described in the previous section (i.e. elastic net regression without PCA) in order to support cross-method comparability. However, it is important to note that the independent variables in this elastic net regression step were PCs rather than individual CpG sites (**Additional file 2: Table S4**). **Additional file 4: Fig. S1** shows associations between the first 20 PCs and relevant covariates in GS.

### 2.5 Evaluation of DNAm predictors of CRP

The relative performances of five distinct DNAm predictors of CRP were compared (hereafter also referred to as DNAm CRP). The measures included those derived from elastic net regression (1), Bayesian penalised regression (2) and PCA with elastic net regression (3). The remaining two predictors were derived using EWAS weights from Wielscher *et al*. (4) and our own linear EWAS (5). Details on assessing the longitudinal trajectories of CRP measures are available in **Additional file 3**. Two metrics were used to assess performance. First, Pearson’s correlations between log-transformed CRP and DNAm CRP measures were computed across all test cohorts. Second, incremental r-squared (*R*^2^) modelling was applied only to a selected test cohort, which was the Lothian Birth Cohort 1936. The cohort was selected in order to align with health outcome association testing (see **Section 3.6**). The time-point of Wave 2 (or age 73, N=756) was selected rather than Wave 1 (age 70, N=885) as high-sensitivity CRP was available at the former but not latter time-point. The incremental r-squared (*R*^2^) was calculated by subtracting the *R*^2^ of the full model from that of the null model as shown below:

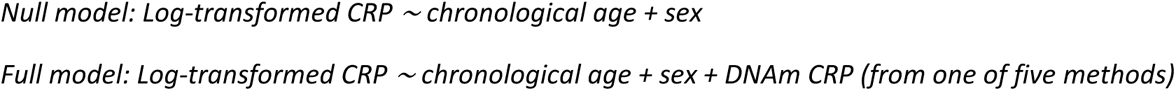

### 2.6 Health outcome association tests

The second Wave of LBC1936 (i.e. at mean age 73 years) was used in health outcome association testing over other cohorts given that the study population was at risk for age-related disease states and frailty phenotypes. The LBC1921 was not used given its smaller analytical sample (N≤170).

Continuous variables were scaled to mean zero and unit variance. Assay-derived CRP or DNAm CRP served as the independent variable in regression models. Linear regression models were used to examine associations between CRP measures and 21 cardiometabolic and fitness variables (dependent variable). Logistic regression tested for associations between CRP measures and lifetime history of four separate conditions (0=’No’, 1=’Yes’). Cox proportional hazard models assessed the relationship between CRP and all-cause mortality. The time-at-risk ran from age at Wave 2 (∼age 73) until the date of recorded death (cases) or the end-of-censor period (controls). All regression models were adjusted for age and sex. Height (in cm) was fitted as an additional fixed-effect covariate for lung function measures and the six metre walk test in linear regression models. A summary of all phenotypes in the LBC1936 is presented in **Additional file 2: Table S5**. Correction for multiple testing was applied using the false discovery rate (FDR *p* < 0.05)^33^.

## 3 Results

### 3.1 Study characteristics and demographics

The six cohorts included in this study had disparate sample sizes and demographic profiles (**Table 1**). The number of individuals included in analyses ranged from 49 (at age 90 in LBC1921) to 17,936 (age range 18-99 years in GS). Mean CRP levels (mg/L) increased across the life-course from 0.2 at birth (cord blood in ALSPAC) to 8.4 at age 90 years (LBC1921).

### 3.2 Epigenome-wide association study to identify individual CpG sites associated with CRP levels

We first investigated marginal associations between log-transformed blood CRP levels and 752,722 CpG sites in GS (N=17,936). There were 33,939 associations with *p* < 3.6×10^-8^ in a basic model that adjusted for chronological age, sex, estimated white blood cell proportions and experimental batch (**Additional file 2: Table S6**). Mixed-effects models that included a kinship matrix were used to account for relatedness as sensitivity analyses^34^. Approximately 75% of associations remained associated (n=25,634 with *p* < 3.6×10^-8^) after accounting for relatedness, and the remainder of associations had *p* < 1×10^-4^. Only 2,805 (8.3%) associations had *p* < 3.6×10^-8^ in a fully-adjusted model that further accounted for five lifestyle factors and population structure (**Fig. 2A, Additional file 2: Table S7**). The substantial attenuation observed after accounting for lifestyle behaviours highlights their associations with DNAm and chronic low-grade inflammation.

**Fig. 2.**
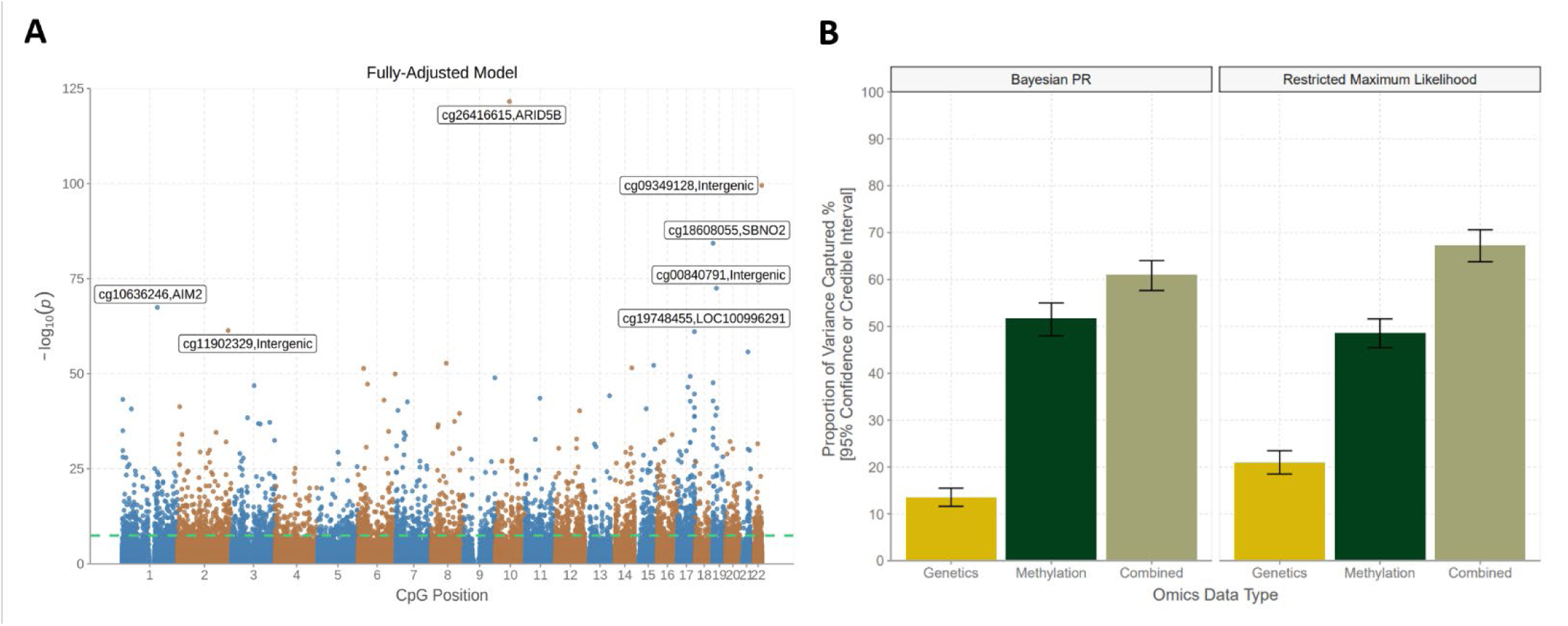
Epigenome-wide association and variance component analyses of blood CRP levels in Generation Scotland. (A) A Manhattan plot shows associations between genome-wide CpG probes and log-transformed CRP levels (N=17,936). Associations from the fully-adjusted model are displayed. The green line denotes the epigenome-wide significance threshold at *p* < 3.6 x 10^-8^. The seven strongest associations (smallest *p*-values) are annotated for clarity. (B) The proportion of variance captured by genome-wide genetic and methylation factors, separately, are shown in gold and dark green bars, respectively. The beige bar details the joint variance captured by genetic and methylation variation when conditioned on one another. Vertical bars denote the 95% credible (Bayesian PR) and confidence (restricted maximum likelihood) intervals, respectively. CpG, cytosine-phosphate-guanine dinucleotide; CRP; C-reactive protein; PR, penalised regression.

A look-up analysis using the EWAS Catalog revealed that 1,496 associations (4.4%) from the basic model were previously reported in the literature^6,7,26,35,36^. Effect sizes for 1,379 significant CpG associations in the largest existing EWAS on CRP were correlated 97% with corresponding associations in our study^6^ (**Additional file 4: Fig. S2**). Lastly, we repeated the basic model using Bayesian penalised regression, which can better account for correlations among probes. This method identified 47 lead CpG sites with a posterior inclusion probability greater than 80%, 38 of which had *p* < 3.6×10^-8^ in the linear model (**see Methods**, **Additional file 2: Table S8**). Our EWAS findings show strong agreement with the existing literature and highlight that a small subset of densely correlated regions show robust associations with CRP levels.

### 3.3 Estimating the proportion of variance in CRP levels captured by global DNAm and genetic factors

Next, we assessed whether global patterns of DNA methylation were associated with individual differences in blood CRP profiles within GS (N=17,936). BayesR+ was used to perform Bayesian penalised regression and Gaussian mixture-based variance partitioning^11^. The proportion of variance captured by genome-wide DNAm alone was 51.7% [95% credible interval (CrI): 48.0%, 55.0%], guiding an upper bound for the performance of DNAm CRP predictors.

Unlike genetic (or SNP-based) heritability estimates, the variance captured by DNAm probes may reflect both cause and consequence on the phenotype. Disentangling the independent and combined contributions from genetic and DNAm variation would refine insights into the molecular architecture of CRP. Using BayesR+, the proportion of variance explained by genetic variation alone was 13.4% [95% CrI: 11.6%, 15.5%], aligning well with a recent SNP-based heritability estimate of 13% from the largest existing GWAS on CRP^37^.

Using BayesR+, the joint variance captured by genetics and DNAm was 61.0% [95% CrI: 57.6%, 64.0%]. The contribution of DNAm to this estimate was 49.0%, which is similar to the estimate from DNAm analysis alone (51.7%) and suggests it was largely independent from underlying genetic factors. Sensitivity analyses were performed in OSCA using a linear mixed model approach with an epigenetics relationship matrix. Estimates from OSCA were highly consistent with those from the Bayesian strategy as illustrated in **Fig. 2B** (**Additional file 2: Table S9**).

### 3.4 Comparing feature selection methods in developing DNAm predictors of CRP

We focused on five distinct methods to generate DNAm predictors of CRP. We trained three predictors using (1) elastic net regression, (2) Bayesian penalised regression and (3) PCA with elastic net regression (‘PCA+elnet’) (N=17,936). We also derived two additional predictors using EWAS weights from (4) Wielscher *et al*. and (5) our own linear EWAS. We then projected them into ALSPAC, HELIOS, SABRE, LBC1936 and LBC1921 (summary data presented in **Additional file 2: Table S10** and **Additional file 5**).

We first assessed the longitudinal stabilities of assay-measured CRP and DNAm CRP (all five predictors) in the LBC1936 (**Additional file 3**). Longitudinal analyses were restricted to Waves 2, 3 and 4 as high-sensitivity CRP measures were only present at these time-points, which enabled consistent comparisons. DNAm predictors showed higher intra-class correlation coefficients (0.84-0.94) than assay-measured CRP (0.78), indicating greater stabilities (**Additional file 2: Table S11**). Linear mixed models were then used to assess whether assay-measured and DNAm CRP derived at Wave 2 showed longitudinal associations with CRP levels over subsequent Waves (i.e. in effect predicting CRP profiles). Assay-measured CRP showed the strongest association with repeat measurements (interaction term between Wave 2 CRP and age: β=−0.20, *p* < 1.6 x 10^-38^) (**Additional file 2: Table S12, Additional file 3**). The elastic net-, Bayesian- and ‘PCA+elnet’-based predictors also exhibited significant albeit weaker associations (β=-0.05 for all, range of *p*=[3.9 x 10^-3^, 7.7 x 10^-3^]). Neither EWAS-based predictor showed a strong association (**Additional file 2: Table 12**). The longitudinal decline in CRP levels in the LBC1936 is apparent in **Table 1** and may reflect attrition bias.

Next, we evaluated correlations between DNAm CRP and assay-measured CRP in all cohorts to identify the best-performing prediction method. Predictors built using elastic net regression, Bayesian penalised regression and ‘PCA+elnet’ showed comparable Pearson’s correlation coefficients with one another in adult samples. Their correlation coefficients ranged from 0.27 to 0.53, 0.28 to 0.49 and 0.28 to 0.47 across cohorts, respectively (**Fig. 3A**, **Additional file 2: Table S13**). Correlations were consistent between males and females, and across genetically diverse individuals in HELIOS and SABRE. Notably, correlations were weaker in childhood samples from ALSPAC including at age 0 (*r*∼0.07) and at age 9 years (*r*∼0.20). These analyses suggested that DNAm predictors of CRP were robust to differences in sex and ethnicity but less so to age variation.

**Fig. 3.**
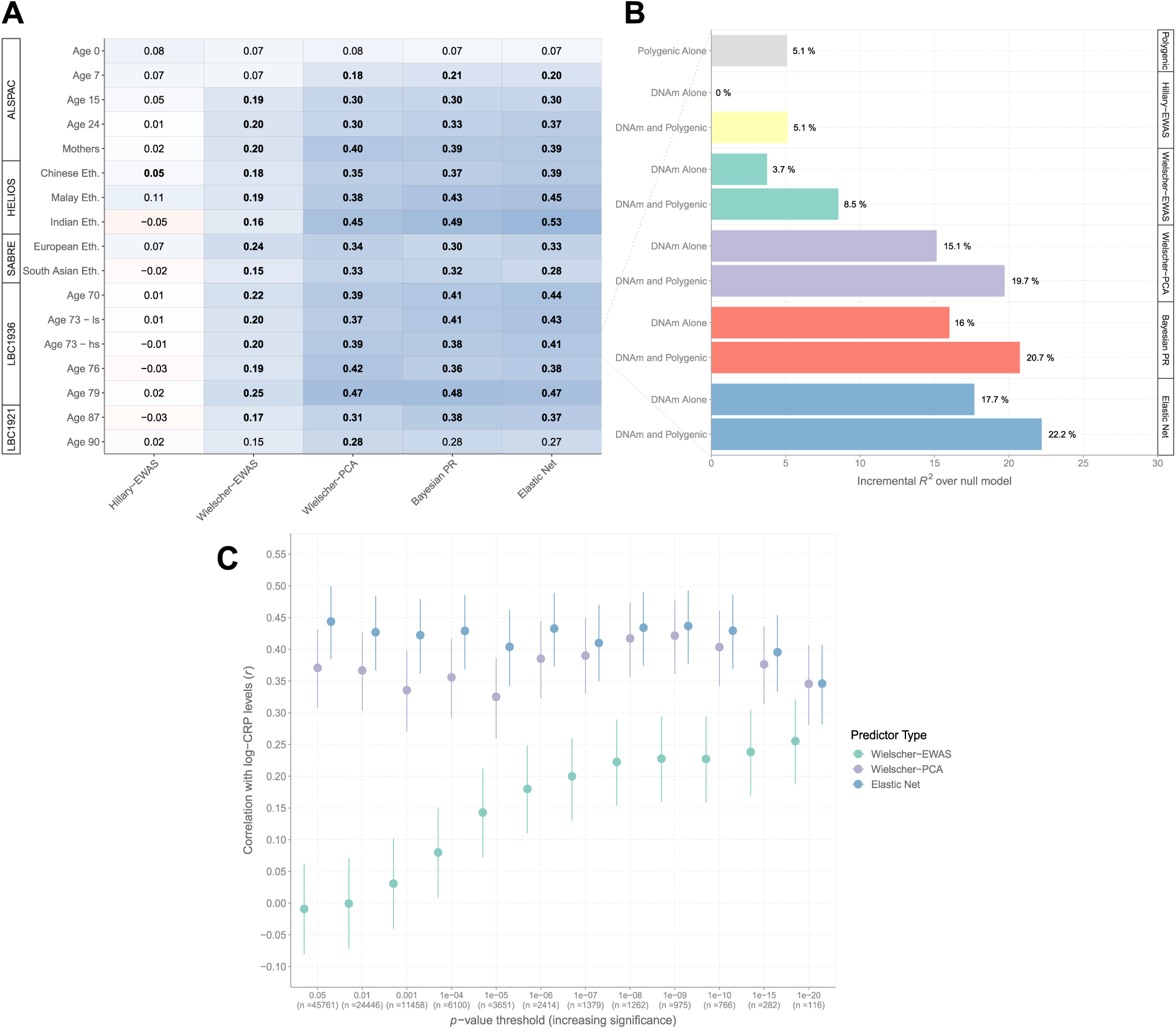
DNAm prediction of blood CRP levels using five separate strategies. (A) Pearson’s correlation coefficients between log-transformed CRP levels and five different DNAm predictors of circulating levels. Weighted linear DNAm predictors for CRP levels were derived from (1) elastic net regression (**Elastic Net**), (2) Bayesian penalised regression (**Bayesian PR**), (3) PCA combined with elastic net regression (**Wielscher-PCA**), (4) an EWAS by Wielscher *et al.* (**Wielscher-EWAS**) and (5) the present EWAS (**Hillary-EWAS**). Low-sensitivity and high-sensitivity CRP measures were available at age 73 (Wave 2) of the LBC1936 and are included in this plot to enable cross-assay comparison. The high-sensitivity measures alone are reported in the main text for this time-point. (B) The proportion of variance captured in log-transformed CRP levels by a polygenic score alone and DNAm CRP from (A) are shown for Wave 2 of the LBC1936 (incremental *R*^2^ estimates above null model, see main text). An additive genetic and DNAm model is also shown for each of the five prediction strategies. (C) The PCA and elastic net regression method in the main text relied on pre-filtering sites to those that surpassed genome-wide significance in the Wielscher *et al*. EWAS (i.e. *p* < 1.0 x 10^-7^). The method was then repeated using different *p*-value thresholds to filter probes prior to PCA. The resulting predictors were compared against (i) weighted linear combinations using EWAS weights alone and (ii) elastic net regression on the filtered CpGs (i.e. bypassing the PCA step). Pearson’s correlations were computed between log-transformed CRP and DNAm CRP for all three methods and for *p*-value thresholds with increasing stringency. Vertical lines denote the 95% confidence interval. ALSPAC, Avon Longitudinal Study of Parents and Children; CpG, cytosine-phosphate-guanine dinucleotide; CRP; C-reactive protein; DNAm, DNA methylation; Eth., Ethnicity; EWAS, epigenome-wide association study; HELIOS, Health for Life in Singapore; LBC1936, Lothian Birth Cohort 1936; PCA, principal component analysis; PR, penalised regression.

The linear predictor – analogous to a polygenic score – based on association effect sizes in the Wielscher EWAS (*p* < 1 x 10^-7^, n≤1,379 sites, **see Methods**) tended to be less correlated with assay-measured CRP (r∼0.2). Retraining the predictor using ‘PCA+elnet’ (and focusing on the same CpG sites) almost doubled the correlations (**Fig. 3A**). There was a negligible correlation between CRP levels and a weighted score derived from our EWAS (n≤33,939 sites with *p* < 3.6×10^-8^), likely reflecting random noise from many more weakly associated individual sites.

The same patterns held true for incremental *R*^2^ estimates of CRP variance explained beyond age and sex. A predictor from elastic net regression explained 17.7% of the variance in CRP over age and sex, with comparable estimates from ‘PCA+elnet’ and Bayesian approaches (**Fig. 3B**). EWAS-based predictors explained only 0-4% of phenotypic variance. A genetic score explained 5.1% of the variance in CRP. DNAm predictors captured variance in CRP independently from the genetic predictor, consistent with the variance partitioning analyses.

### 3.5 Elastic net regression is sufficient to enhance DNAm prediction of CRP

The primary ‘PCA+elnet’-based approach was applied to 1,379 sites that associated with CRP at *p* < 1.0 x 10^-7^ in the Wielscher *et al.* study. The strategy was repeated across a range of pre-filtering *p*-value thresholds to determine its sensitivity to specific thresholds (*p* < 0.05 as the least stringent to *p* < 1 x 10^-20^ as the most stringent). The ‘PCA+elnet’ predictor performed similarly regardless of the threshold (*r*=0.33-0.42). We also trained the predictor using sites common to all cohorts (n=374,785) to directly match the original strategy used by Higgins Chen *et al.*^12^. The resulting correlation coefficient was slightly lower (r=0.30).

By contrast, the performance of the EWAS-based predictor tended to improve at more stringent *p*-value thresholds. The correlation coefficient ranged from −0.01 (using sites at *p* < 0.05) to 0.26 (*p* < 1 x 10^-20^) (**Fig. 3C, Additional file 2: Table S14**).

Additionally, we repeated the approach without the PCA step and retrained predictors using elastic net regression alone. Elastic net alone outperformed the combined ‘PCA+elnet’ strategy, on average, by 10%, suggesting that the PCA step did not enhance predictive performance.

### 3.6 Comparing DNAm and assay-measured CRP in their associations with cardiometabolic health outcomes

Lastly, we compared DNAm CRP and assay-measured CRP in their associations with 26 health outcomes at age 73 years in the LBC1936 (**see Methods**). Previous studies have focused on EWAS-based predictors alone. Here, we focused on the elastic net-based predictor given that it was the best-performing method in correlation analyses and incremental *R*^2^ modelling. Overall, assay-measured CRP and DNAm CRP displayed similar relationships with continuous outcomes with 15 and 13 associations (from 21 outcomes) having *p_FDR_* < 0.05, respectively (**Fig. 4**, **Additional file 2: Table S15**).

**Fig. 4.**
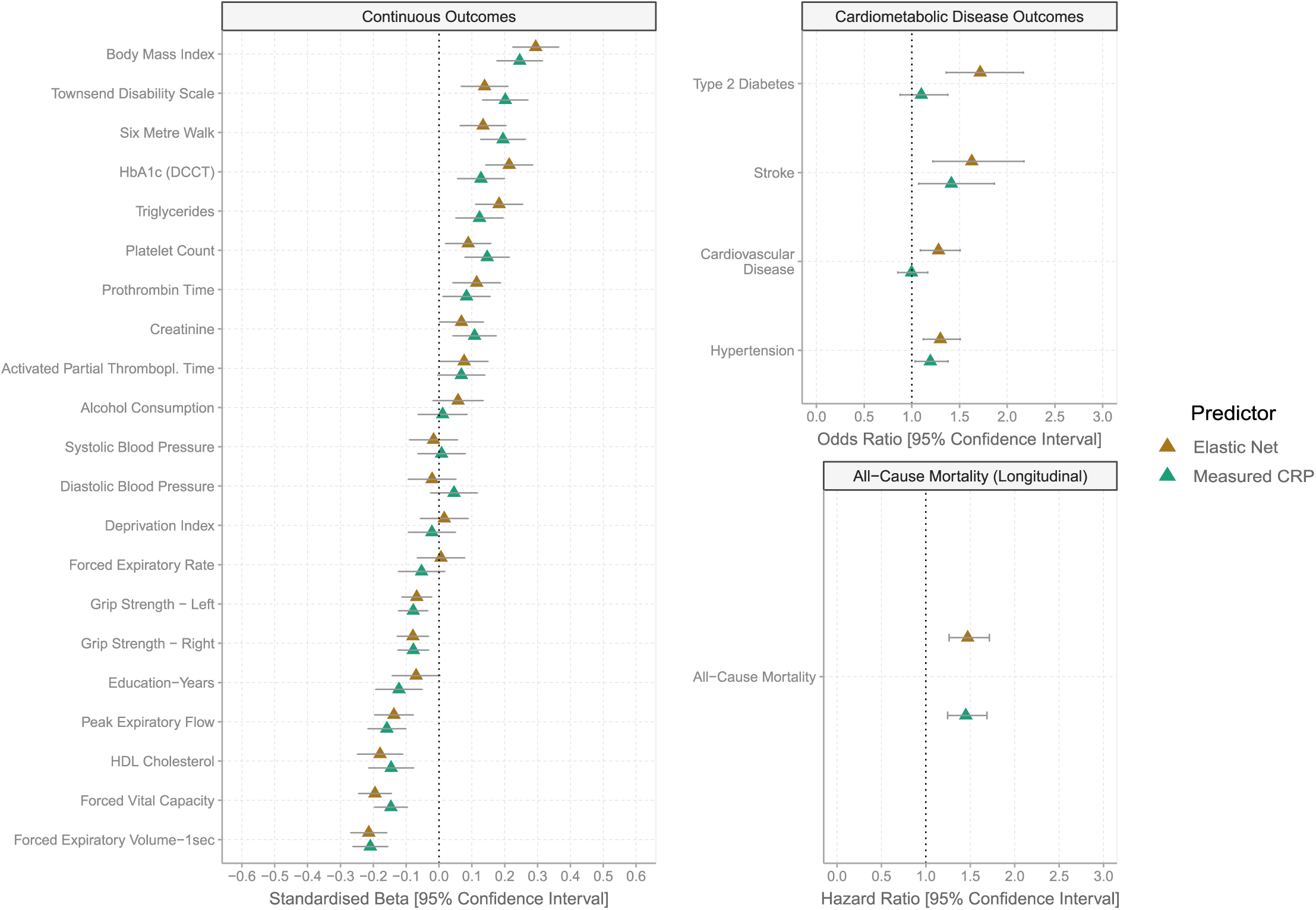
Associations of health outcomes with DNAm CRP from elastic net regression and phenotypic CRP. Linear and logistic regression models were used to test for cross-sectional associations of DNAm and assay-measured (i.e. phenotypic) CRP with cardiometabolic, lifestyle and self-report disease variables at Wave 2 of the LBC1936. Cox proportional hazard models tested for associations between CRP (assay-measured or DNAm) derived at Wave 2 and time-to-death due to all-cause mortality. Here, only DNAm CRP from elastic net regression was utilised given that it was deemed the best-performing method in correlation analysis and incremental *R*^2^ modelling. Association tests using DNAm CRP from other prediction strategies and a polygenic score for CRP are shown in **Additional file 4: Fig. S3-S8**. CRP; C-reactive protein; DCCT; Diabetes Control and Complications Trial; DNAm, DNA methylation; HDL, high-density lipoprotein; Thrombopl., thromboplastin.

However, for cardiometabolic disease outcomes, DNAm CRP outperformed assay-measured CRP (**Fig. 4**). Whereas DNAm CRP was strongly associated with history of cardiovascular disease (odds ratio (OR)=1.28, *p*_FDR_=6.6 x 10^-3^), hypertension (OR=1.30, *p*_FDR_=2.3 x 10^-3^), stroke (OR=1.63, *p*_FDR_=3.4 x 10^-3^) and type 2 diabetes (OR=1.72, *p*_FDR_=8.0 x 10^-5^), assay-measured CRP was weakly associated only with hypertension (OR=1.19, *p*_FDR_=0.03) and stroke (OR=1.41, *p*_FDR_=0.03). Both measures were similarly associated with risk of all-cause mortality (hazard ratios=1.47 and 1.45, *p*_FDR_=3.0 x 10^-6^ and 3.7 x 10^-6^, respectively, **Fig. 4**).

Association patterns were similar for the Bayesian- and ‘PCA+elnet’-based predictors, which associated with 19 and 20 outcomes at *p*_FDR_ < 0.05, respectively. The EWAS-based predictors associated with 8 (our EWAS) and 15 outcomes (Wielscher EWAS) (**Additional file 4: Fig. S3-S8**). Associations for the genetic score did not withstand multiple testing correction. Therefore, our newly-described DNAm models capture adverse health outcomes better than assay-measured CRP as well as existing DNAm and genetic models of CRP.

## 4 Discussion

We developed a DNAm predictor of CRP that explains up to 20% of the variance in circulating concentrations, almost doubling that captured by existing predictors^7,8^. DNAm CRP also outperforms genetic scores and assay-measured CRP in association analyses with cardiometabolic disease outcomes and risk factors. While the DNAm predictor was developed using data from over 17,000 Scottish adults, it is generalisable across cohorts with distinct birth periods, DNAm normalisation pipelines, sex differences and individuals of different ethnic backgrounds. These analyses comprehensively outline the utility of DNAm as a biomarker of chronic low-grade inflammation and provide new opportunities to capture inflammatory burden across diverse populations.

We identified elastic net regression as the best-performing method to proxy chronic low-grade inflammation from DNAm. Recently, Higgins Chen *et al.* showed that PCA prior to elastic net regression improved the reliability of epigenetic age estimators^12^. Similarly, Doherty *et al.* found that PCA in advance of elastic net regression outperformed 12 other strategies including elastic net regression alone in the DNAm prediction of telomere length^38^. Trejo-Banos *et al.* showed that DNAm predictors of BMI and cigarette smoking developed using Bayesian penalised regression captured more phenotypic variance than those from conventional penalised regression methods^11,39^. Here, elastic net regression without PCA offered a slight increase in performance over these methods (∼1-2%) in explaining CRP variance. We show that this benefit holds over a range of pre-filtering criteria. The method also offers greater interpretability in having fewer selected features and lower computational expense than the PCA-based approach. Furthermore, it provides faster run times than BayesR+ (minutes versus hours based on current software versions). Nevertheless, the optimal method for a given trait is likely to depend on the precise relationship between the phenotype and molecular dataset in question.

Existing DNAm predictors of inflammation represent additive weighted scores that consider CpG sites and weights (i.e. coefficients) from EWAS alone. This approach is analogous to the development of polygenic scores. Verschoor *et al.* showed that an aggregate score derived from the recent EWAS by Wielscher *et al.* outperformed assay-measured CRP as a marker of cardiopulmonary disease and long-term health status. However, neither measure associated with all-cause mortality, which is in contrast with our study^10^. A seven-CpG score derived from an earlier EWAS by Ligthart *et al.* has also been associated with a wide range of neurocognitive health outcomes in adults and neonates^7–9,40–42^. These aggregate EWAS scores have shown promise in correlating with cardiometabolic disease risk and risk factors^6,7^. However, we show that predictors or measures from feature selection methods (i.e. elastic net regression) capture much more trait variance and associate with a greater number of outcomes. Our newly-described predictors performed comparably in residents of the UK and Singapore with diverse self-report ethnicities, which indicates its potential as a biomarker of inflammation in different populations. However, there remains a need to expand prediction efforts to individuals across other global regions and of additional ethnic backgrounds and ancestries in order to capture a fuller range of genetic and environmental contexts. Our predictors were developed using whole-blood adult samples but performed poorly at the extremes of the life-course including in neonates and in the ninth decade of life. Indeed, the predictors had near-zero correlations in cord blood samples. Cord blood DNAm serves a better proxy for cord blood CRP than maternal blood DNAm^43^. Therefore, additional tissue sources may be required to improve the generalisability of inflammatory biomarkers alongside environmental, technical and statistical considerations.

The strengths of this study include the use of a diverse range of cohorts that span the life-course and allow for a comprehensive examination of generalisability. We also utilise multiple methods to inform the molecular architecture of circulating CRP levels and provide robust estimates for the contribution of genome-wide DNAm to inter-individual variability. Limitations of the study include the non-consideration of medication data and cross-sectional measurements preventing evaluation of the capacity of CRP models to predict health outcomes. The non-consideration of medication data and the advanced age of the LBC1936 cohort could have influenced associations between measured CRP and cross-sectional health outcomes. Additional limitations include different platforms for methylation typing and CRP measurements, small numbers of analytical samples in some cohorts (e.g. LBC1921) and the potential for non-linear associations between CRP and CpG methylation. Future work should focus on considering further advanced statistical methodologies that can consider nuanced and complex relationships between DNAm and CRP in addition to other human traits.

## 5 Conclusion

Genome-wide DNAm serves as a strong proxy for chronic low-grade inflammation, capturing up to 50% of inter-individual variation in circulating CRP levels. Our newly-described DNAm predictors of CRP are generalisable across the population cohorts tested and offer new and improved opportunities to examine the association between chronic inflammation and health outcomes across disparate clinical and regional populations.

## Data Availability

According to the terms of consent for GS participants, access to data must be reviewed by the GS Access Committee. Applications should be made to. ALSPAC data access is through a system of managed open access. Submissions and queries should be directed to. For HELIOS, data access request proposals should be directed to for the consideration of the HELIOS Studys principal investigators. SABRE data used for this submission will be made available on request to. Further details regarding data sharing can be found on the cohort web pages (https://www.sabrestudy.org/home-2/data-sharing/). Lothian Birth Cohort data access requests can be made by following the information at https://www.ed.ac.uk/lothian-birth-cohorts/data-access-collaboration.
Epigenome–wide association statistics from linear models will be made available at the EWAS Catalog [URL to follow at publication]. Epigenome–wide association statistics from Bayesian penalised regression will be made available at the University of Edinburgh Datashare site [URL to follow at publication]. CpGs and weights derived from elastic net regression, Bayesian penalised regression and the combined PCA and elastic net regression strategies will also be available at the Edinburgh Datashare site [URL to follow at publication]. CpGs and weights for the elastic net regression and PCA+elnet–based approaches from this version of the manuscript are available in Additional file 2: Tables S3 and S4. The Bayesian penalised regression predictor is not made available in Additional file 2 due to the large number of CpGs involved (>300,000). All code associated with this manuscript, including scripts to project DNAm CRP into test samples, is available open access at the following GitHub repository: https://github.com/robertfhillary/dnam-crp.

## Acknowledgements

**This research was funded in whole, or in part, by Wellcome [104036/Z/14/Z, 220857/Z/20/Z, 217065/Z/19/Z, 067100, 37055891, 086676/7/08/Z, 221890/Z/20/Z]. For the purpose of open access, the author has applied a CC BY public copyright licence to any Author Accepted Manuscript version arising from this submission.**

We are extremely grateful to all participants, team members and support staff in GS, HELIOS, SABRE and the LBCs for their invaluable contributions to this study. Further, for ALSPAC, we are extremely grateful to all the families who took part in this study, the midwives for their help in recruiting them, and the whole ALSPAC team, which includes interviewers, computer and laboratory technicians, clerical workers, research scientists, volunteers, managers, receptionists and nurses.

GS received core support from the Chief Scientist Office of the Scottish Government Health Directorates [**CZD/16/6**] and the Scottish Funding Council [**HR03006**]. Genotyping and DNA methylation profiling of the GS samples was carried out by the Genetics Core Laboratory at the Edinburgh Clinical Research Facility, Edinburgh, Scotland, and was funded by the Medical Research Council UK and Wellcome (Wellcome Trust Strategic Award STratifying Resilience and Depression Longitudinally [STRADL; Reference **104036/Z/14/Z**]). The DNA methylation data assayed for GS was partially funded by Wellcome [**220857/Z/20/Z**], a 2018 NARSAD Young Investigator Grant from the Brain & Behavior Research Foundation [**27404**; awardee: Dr David M Howard] and by a JMAS SIM fellowship from the Royal College of Physicians of Edinburgh [Awardee: Dr Heather C Whalley]. Roche Diagnostics supported this study through provision of free reagents and a grant for measurement of CRP in GS. We thank Elaine Butler, Ross Hepburn, and Ellen Macdonald, University of Glasgow, for excellent technical support. The UK Medical Research Council and Wellcome [**217065/Z/19/Z**] and the University of Bristol provided core support for ALSPAC. A comprehensive list of grants funding is available on the ALSPAC website (http://www.bristol.ac.uk/alspac/external/documents/grant-acknowledgements.pdf). Methylation data in the ALSPAC cohort were generated as part of the UK BBSRC-funded [**BB/I025751/1**, **BB/I025263/1**] Accessible Resource for Integrated Epigenomic Studies (ARIES, https://www.ariesepigenomics.org.uk). The ALSPAC study was further supported by the National Institute for Health and Care Research Bristol Biomedical Research Centre. The views expressed are those of the author(s) and not necessarily those of the NIHR or the Department of Health and Social Care. The HELIOS study was supported by the Singapore Ministry of Health’s National Medical Research Council under its OF-LCG funding scheme [**MOH-000271-00**] and intramural funding from Nanyang Technological University, Lee Kong Chian School of Medicine and the National Healthcare Group of Singapore. SABRE was supported at baseline by the Medical Research Council, the British Heart Foundation and Diabetes UK. At follow-up, the SABRE study was funded by Wellcome [**067100**, **37055891**, **086676/7/08/Z**], the British Heart Foundation [**PG/06/145**, **PG/08/103/26133**, **PG/12/ 29/29497**, **CS/13/1/30327**] and Diabetes UK [**13/0004774**]. The SABRE study team also acknowledges the support of the National Institute of Health Research Clinical Research Network [**NIHRCRN**]. The LBC1936 is jointly core-funded by the Biotechnology and Biological Sciences Research Council and the Economic and Social Research Council [**BB/W008793/1**], and received support from Age UK (Disconnected Mind programme), the Milton Damerel Trust, the Medical Research Council [**MR/M01311/1**] and the University of Edinburgh. LBC1921 data collection was supported by grants from the Biotechnology and Biological Sciences Research Council [**15/SAG09977**] and the Chief Scientist Office of the Scottish Executive Health Department [**CZB/4/505, ETM/55, CZH/4/213, CZG/3/2/79**]. Methylation typing was supported by the Centre for Cognitive Ageing and Cognitive Epidemiology (Pilot Fund award), Age UK, The Wellcome Trust Institutional Strategic Support Fund, The University of Edinburgh, and The University of Queensland.

R.F.H. is supported by a British Heart Foundation Immediate Fellowship [**FS/IPBSRF/22/27042**]. H.R.E. is supported by the Medical Research Council Integrative Epidemiology Unit at the University of Bristol [**MC_UU_00011/5**]. F.H. and K.D. were supported within a Unit that received support from the UK Medical Research Council [**MC_UU_12019/1**]. S.R.C. was supported by a Sir Henry Dale Fellowship jointly funded by Wellcome and the Royal Society [**221890/Z/20/Z**]. R.E.M. is supported by an Alzheimer’s Society major project grant [**AS-PG-19b-010**]. P.D.Y. and M.S. are supported by the Medical Research Council Integrative Epidemiology Unit at the University of Bristol [**MC_UU_00011/5**] and Cancer Research UK [**C18281/A29019**].

## Author contributions

R.F.H., C.L.R., R.E.M., P.D.Y. and M.S. conceptualised the study design. R.F.H., H.K.N., P.D.Y. and M.S. performed the analyses. D.L.Mc.C., H.R.E., R.M.W., A.C., F.H., D.K., P.W., N.S., J.C., C.H., C.S., A.M.M., K.L.E, S.R.C., J.C.C. and M.L. were involved in data generation and preparation. All authors reviewed and approved of the manuscript.

## Ethics statements

### Generation Scotland

All components of Generation Scotland received ethical approval from the NHS Tayside Committee on Medical Research Ethics [REC Reference Number: **05/S1401/89**]. Generation Scotland has also been granted Research Tissue Bank status by the East of Scotland Research Ethics Service [REC Reference Number: **20-ES-0021**], providing generic ethical approval for a wide range of uses within medical research. All participants provided written informed consent.

### ALSPAC

Ethical approval for the study was obtained from the ALSPAC Ethics and Law Committee and the Local Research Ethics Committees. Informed consent for the use of data collected via questionnaires and clinics was obtained from participants following the recommendations of the ALSPAC Ethics and Law Committee at the time. Consent for biological samples has been collected in accordance with the Human Tissue Act (2004).

### HELIOS

The HELIOS study was approved by the National Technological University (NTU) Institutional Review Board [**IRB-2016-11-030**], with written informed consent obtained from each participant before the commencement of the study.

### SABRE

The SABRE study was approved by St Mary’s Hospital Research Ethics Committee [**07/H0712/109**] and all participants provided written informed consent.

### Lothian Birth Cohort 1936

Ethical approval was obtained from the Multicentre Research Ethics Committee for Scotland (age 70, [**MREC/01/0/56**]), the Lothian Research Ethics Committee (age 70, [**LREC/2003/2/29**]), and the Scotland A Research Ethics Committee (ages 73, 76, 79, 82, [**07/MRE00/58**]). All participants provided written informed consent.

### Lothian Birth Cohort 1921

Ethical approval was provided by the Lothian Research Ethics Committee for test waves 1–3 at ages 79, 83 and 87 [**LREC/1998/4/183, LREC/2003/7/23, 1702/98/4/183**] and the Scotland A Research Ethics Committee for test wave 4 at age 90 [**10/MRE00/87, 10/MRE00/87**]. All participants provided written informed consent.

## Availability of data and materials

According to the terms of consent for GS participants, access to data must be reviewed by the GS Access Committee. Applications should be made to. ALSPAC data access is through a system of managed open access. Submissions and queries should be directed to. For HELIOS, data access request proposals should be directed to for the consideration of the HELIOS Study’s principal investigators. SABRE data used for this submission will be made available on request to. Further details regarding data sharing can be found on the cohort web pages (https://www.sabrestudy.org/home-2/data-sharing/). Lothian Birth Cohort data access requests can be made by following the information at https://www.ed.ac.uk/lothian-birth-cohorts/data-access-collaboration.

Epigenome-wide association statistics from linear models will be made available at the EWAS Catalog [URL to follow at publication]. Epigenome-wide association statistics from Bayesian penalised regression will be made available at the University of Edinburgh Datashare site [URL to follow at publication]. CpGs and weights derived from elastic net regression, Bayesian penalised regression and the combined PCA and elastic net regression strategies will also be available at the Edinburgh Datashare site [URL to follow at publication]. CpGs and weights for the elastic net regression and ‘PCA+elnet’-based approaches from this version of the manuscript are available in **Additional file 2: Tables S3 and S4.** The Bayesian penalised regression predictor is not made available in **Additional file 2** due to the large number of CpGs involved (>300,000). All code associated with this manuscript, including scripts to project DNAm CRP into test samples, is available open access at the following GitHub repository: https://github.com/robertfhillary/dnam-crp.

## Supplementary information captions

- **Additional file 1 – Cohort Descriptions.**
- **Additional file 2 - Supplementary Tables. Table S1**. Summary data for Generation Scotland data. **Table S2**. Associations between blood CRP and covariates in Generation Scotland. **Table S3**. CpGs and weights for elastic net predictor. **Table S4**. CpGs and weights for PCA-based predictor. **Table S5**. Summary data for LBC1936 phenotypes. **Table S6**. Results from basic model - epigenome-wide association study. **Table S7**. Results from fully-adjusted model - epigenome-wide association study. **Table S8**. Results from Bayesian penalised regression - epigenome-wide association study. **Table S9**. Variance components estimation. **Table S10**. Summary data for DNAm CRP predictors across cohorts. **Table S11**. Intra-class correlation coefficients for DNAm and assay-measured CRP over three waves of the LBC1936. **Table S12**. Longitudinal associations between CRP measures at Wave 2 and assay-measured CRP in the LBC1936. **Table S13**. Correlation between DNAm CRP and phenotypic CRP using five strategies. **Table S14**. Influence of pre-filtering threshold on the performance of DNAm prediction. **Table S15**. Health outcome association testing in the LBC1936.
- **Additional file 3 – Supplementary Methods**. Additional descriptions on (i) Bayesian penalised regression, (ii) variance component estimation – combined analysis, (iii) assessing longitudinal stability of CRP measures, (iv) assessing relationship between assay-measured or DNAm CRP and future CRP measurements, (v) polygenic score profiling in Lothian Birth Cohort 1936 and (vi) note on elastic net regression.
- **Additional file 4 - Supplementary Figures. Fig. S1**. Correlation between the first 20 principal components on CRP-associated probes and continuous covariates in Generation Scotland. **Fig. S2**. Correlation of effect sizes between epigenome-wide association studies by *Wielscher et al.* and the present study (Hillary *et al.*). **Fig. S3**. Association of 21 continuous cardiometabolic and lifestyle variables with measured CRP, genetic score for CRP and five DNAm predictors of CRP. **Fig. S4**. Associations of health outcomes with a genetic score for CRP or assay-measured CRP in the Lothian Birth Cohort 1936. **Fig. S5**. Associations of health outcomes with DNAm CRP (Hillary EWAS-based predictor) or assay-measured CRP in the Lothian Birth Cohort 1936. **Fig. S6**. Associations of health outcomes with DNAm CRP (Wielscher EWAS-based predictor) or assay-measured CRP in the Lothian Birth Cohort 1936. **Fig. S7**. Associations of health outcomes with DNAm CRP (PCA-based predictor) or assay-measured CRP in the Lothian Birth Cohort 1936. **Fig. S8**. Associations of health outcomes with DNAm CRP (Bayesian PR-based predictor) or assay-measured CRP in the Lothian Birth Cohort 1936.
- **Additional file 5 - Correlations between DNAm predictors within each cohort and sub-group.**

