## Additional file 1 for "Blood-based epigenome-wide analyses of chronic low-grade inflammation across diverse population cohorts"

**Additional file 1 – Cohort Descriptions.** The following information pertains to Cohort Descriptions for the manuscript ‘*Blood-based epigenome-wide analyses of chronic low-grade inflammation across diverse population cohorts’* by Hillary *et al.*

**Training Cohort: Generation Scotland**

*DNA methylation in Generation Scotland*

Whole-blood DNAm was measured using the Illumina Infinium MethylationEPIC array. DNAm was assayed in three distinct sets (*N*_Set1_ = 5,087, *N*_Set2_ = 4,450, *N*_Set3_ = 8,876) and 121 experimental batches. Set 1 contained related individuals. Set 2 consisted of individuals who were unrelated to each other and those in Set 1. Set 3 consisted of related individuals, and individuals related to those in Sets 1 and 2. Set 1 followed a slightly different quality control strategy to Sets 2 and 3, which together followed the same strategy. In Set 1, samples were removed if: (i) ≥1% of probes had a detection *p*-value >0.05 or (ii) there was a disagreement between self-reported sex and methylation-predicted sex. Probes were removed if: (i) ≥5% of samples had a bead count <3 or a detection *p*-value >0.05, (ii) they were non-autosomal or (iii) they overlay any SNPs and/or resided in potential cross-hybridising locations. In Sets 2 and 3, samples were removed if: (i) ≥0.5% of probes had a detection *p*-value >0.01 or (ii) there was a disagreement between self-reported sex and methylation-predicted sex. Probes were excluded if (i) ≥5% of samples had a bead count of 3 or less, (ii) ≥1% of samples had a detection *p*-value >0.01, (iii) they were non-autosomal or (iii) they overlay any SNPs and/or resided in potential cross-hybridising locations. There were 752,722 probes and 18,413 individuals with DNA methylation measurements following quality control.

*CRP measurements in Generation Scotland*

Serum CRP levels (mg/L) were quantified at the University of Glasgow using a commercial high-sensitivity assay on an automated analyser (c311, Roche Diagnostics, UK). Manufacturer’s calibration and quality control were employed. In total, 17,936 individuals had paired genome-wide DNAm and CRP measurements following quality control.

**Test Cohorts**

**Avon Longitudinal Study of Children and Parents**

*Cohort description*

Pregnant women resident in Avon, UK with expected dates of delivery between 1st April 1991 and 31st December 1992 were invited to take part in the study. The initial number of pregnancies enrolled was 14,541 leading to 13,988 children who were alive at 1 year of age. As part of Accessible Resource for Integrated Epigenomic Studies (ARIES)^1^, a sub-sample of 1,018 ALSPAC mother-child pairs had blood DNAm assayed using the Illumina 450 K array. DNAm was assayed in peripheral blood at all time-points except for age 0 (cord blood) and age 9 (B-cell-enriched buffy coat). High-sensitivity CRP was quantified using an automated particle-enhanced immunoturbidimetric assay (Roche UK, Welwyn Garden City, UK). CRP was measured in peripheral blood at all time-points with the exception of cord blood.

Study data were collected and managed using REDCap electronic data capture tools hosted at the University of Bristol. REDCap (Research Electronic Data Capture) is a secure, web-based software platform designed to support data capture for research studies^2^. Please note that the study website contains details of all the data that is available through a fully searchable data dictionary and variable search tool (http://www.bristol.ac.uk/alspac/researchers/our-data/).

*DNA methylation in ALSPAC*

The Illumina 450 K methylation array was used to assess genome-wide DNAm patterns. There were four measurements available for the children involved the cohort. These time-points were age 0, age 9, age 15 or 17 and age 24. DNA methylation was also assessed in the mothers involved in the cohort 18 years after recruitment and the study pregnancy. Therefore, five separate time-points are represented in the analyses. DNAm was measured in cord blood (age 0), peripheral blood (age 15 or 17, 24, and the mothers’ samples) and B-cell-enriched buffy coat samples at age 9. Samples across different time-points were distributed in a semi-random manner across slides in order to mitigate batch effects. Data pre-processing was performed using the R package *Meffil*. Samples that had an average probe detection *p* ≥ 0.01 were removed, along with those that had sex or genotype mismatches. Probes with detection *p*-values < 0.01 were excluded. There were 483,068 probes.

*CRP measurement in ALSPAC*

High-sensitivity CRP was quantified using an automated particle-enhanced immunoturbidimetric assay (Roche UK, Welwyn Garden City, UK). CRP was either measured in cord blood (age 0) or peripheral blood at all other time-points. All assay coefficients of variation were <5%. Following quality control, there were 389, 336, 945, 745 and 773 samples corresponding to age 0, age 9, age 15 or 17, age 24 and the mothers, respectively.

**Health for Life in Singapore**

*Cohort description*

HELIOS is a population-based cohort comprising 10,000 Asian men and women living in Singapore^3^. The cohort aims to recruit 100,000 individuals within Singapore. HELIOS includes Singapore citizens or Permanent Residents aged 30–84 years old. Participants were recruited from the general population and through community outreach programs to ensure diversity in ethnicity and socio-economic backgrounds.

*DNA methylation in HELIOS*

Methylation of genomic DNA was initially quantified in 2,400 samples using the Illumina MethylationEPIC array according to manufacturer’s instructions. Bisulfite conversion of genomic DNA was performed using the EZ DNA methylation kit according to manufacturer's instructions (Zymo Research, Orange, CA). Bead intensity was retrieved using the *minfi* software package, and a detection *p*-value of <0.01 was used for marker calling. In total, 846,604 positions were assayed on the array. Markers with call rates beneath 95% were removed (n=8,882). Fifty-eight samples were excluded; two for array scanning failure, 39 for gender inconsistency and 17 duplicates. None of the samples failed the sample call rate criterion (<95%). Marker intensities were normalised by quantile normalisation^4,5^. This left 837,722 CpG sites for analyses.

*CRP measurement in HELIOS*

CRP levels were measured using wide-range CRP technology, which has been proposed as an economic alternative to high-sensitivity CRP screening^6^. C-reactive protein (CRP) was measured from fasting blood samples by the accredited laboratory (QuestLab, Singapore, SAC–SINGLAS ISO 15189:2012) using ADVIA 1800 chemistry system (Siemens Healthcare, Munich, Germany). Following quality control and outlier removal, there were 1,778, 242 and 225 samples corresponding to the Chinese, Malay and Indian ethnicity groups within HELIOS, respectively.

**Southall And Brent REvisited**

*Cohort description*

The cohort was formed from two identical cross-sectional studies, conducted by the same team, based in the London boroughs of Southall and Brent. Participants were aged between 40 and 69 at the study baseline. Whole-blood DNAm was measured using the Illumina 450 K methylation array in SABRE. High sensitivity CRP was measured using an automated platform (c311 Roche Diagnostics, Burgess Hill UK).

*DNA methylation in SABRE*

Whole-blood DNAm was measured using the Illumina 450 K methylation array in SABRE. Quality control was performed using the *meffil.qc.parameters* function in the R package *Meffil*^7^. Baseline and follow-up samples (derived approximately 20 years later) were analysed together. The following amendments were made to the function’s default pipeline: probes and samples with a detection *p*-value < 0.1 were excluded, the bead number threshold was reduced from 0.2 to 0.1, sample genotype concordance was reduced from 0.9 to 0.8 and the standard deviation (SD) multiple at which sex outliers are identified was raised from 3 to 5 SDs. Eleven samples and 1,644 probes did not meet quality control criteria. The final dataset included 1,422 baseline samples, 589 follow-up samples and 484,781 probes.

*CRP measurement in SABRE*

High sensitivity CRP was measured using an automated platform (c311 Roche Diagnostics, Burgess Hill UK). CRP measurements were available for follow-up samples. There were 588 individuals with paired genome-wide DNAm and CRP available at the follow-up time-point. Of these, 315 and 273 were of European and South Asian ancestry, respectively.

**Lothian Birth Cohorts of 1921 and LBC1936**

*Cohort description*

Participants were born in 1921 or 1936 in the respective studies and completed an intelligence test at age 11 years (in 1932 or 1947). Participants who were living within Edinburgh and the surrounding Lothian regions were re-contacted and entered the study at mean ages of 70 years (LBC1936) and 79 years (LBC1921). The participants have been followed up approximately every three years for a series of clinical, physical, cognitive, biological and sociodemographic data collection. Baseline is denoted as Wave 1 within each cohort. Whole-blood DNAm was assayed using the Illumina 450 K methylation array. CRP levels (mg/L) were quantified using both a high-sensitivity (ELISA; R&D Systems) and a low-sensitivity assay with a dry-slide immuno-rate method on an OrthoFusion 5.1 F.S analyser (Ortho Clinical Diagnostics). Paired DNAm and CRP data were available at Waves 1-4 for the LBC1936 (age 70-79 years) and Waves 3 and 4 for the LBC1921 (age 87 and 90 years).

*DNA methylation in LBC1921 and LBC1936*

DNA from whole-blood was assayed using the Illumina 450 K methylation array. Raw intensity data were background-corrected and normalised using internal controls^8,9^. Methylation was assayed at three time separate points (Set 1, Set 2 and Set 3). The three sets included 2,195, 996 and 552 samples, respectively. Each set began with 485,512 CpGs. Twenty-three duplicate samples were removed from Set 2. Set 1 and Set 2 had 123 duplicates between them, and a sample was removed from each duplicate pair (108 from Set 1 and 15 from Set 2). Sets 1 and 2 were then combined (as Set1-2). Ten duplicate samples were excluded from Set 3. There were also 31 duplicates between Set 3 and the newly-formed Set 1-2. Twenty-six samples were removed from Set 3 and five were removed from Set1-2. The three sets were then combined together (Set1-2-3) and comprised 3,556 samples. Samples and CpGs were filtered on low call rates (CpGs with a detection *p*-value ≥0.01), with a threshold of 95% for both samples and CpGs. In total, 3,525 samples (spread across Waves and LBC1921/LBC1936) and 470,278 CpGs remained. Finally, sex chromosome probes were removed, leaving a dataset that consisted 459,309 CpGs and 3,525 samples.

*CRP measurement in LBC1921 and LBC1936*

Serum CRP was measured from venesected whole-blood samples. CRP levels (mg/L) were quantified using both a high-sensitivity (ELISA; R&D Systems) and a low-sensitivity assay with a dry-slide immuno-rate method on an OrthoFusion 5.1 F.S analyser (Ortho Clinical Diagnostics). The low-sensitivity assay cannot distinguish between values less than 3 mg/L. Therefore, all readings of < 3 mg/L using the low-sensitivity assay were assigned a value of 1.5 mg/L. In the LBC1921, serum CRP was measured via the low-sensitivity method at Waves 3 and 4. The number of individuals with complete paired data was 170 and 49 at these LBC1921 Waves, respectively. In the LBC1936, only low-sensitivity measures were available for Wave 1 (age 70 years) whereas high-sensitivity measures were available for Waves 2, 3, and 4 (ages 73, 76 and 79 years). The final analytical sample contained 885, 756, 536 and 492 individuals at these time-points, respectively.
