## Additional file 3 for "Blood-based epigenome-wide analyses of chronic low-grade inflammation across diverse population cohorts"

**Additional file 3 - Supplementary Methods.** The following information pertains to Supplementary Methods for the manuscript ‘*Blood-based epigenome-wide analyses of chronic low-grade inflammation across diverse population cohorts’* by Hillary *et al.*

**Bayesian penalised regression**

The Gibbs algorithm consisted of 10,000 samples (in other words, iterations) and 5,000 samples of burn-in. A thinning of 5 samples was applied in order to reduce autocorrelation. The process left 1,000 samples and was repeated over four chains, which were initialised using a different random number seed for each chain. The final 250 samples (of 1,000) from each chain were combined for downstream analyses, giving a total of 1,000 samples for our analyses. Penalised regression does not consider the identity of probes, which may preclude biological inference. Indeed, the method may select one member from a correlated group of probes as the representative signal in one iteration and another member in the next, thereby distributing their inclusion probabilities. For the epigenome-wide association study (EWAS), CpGs within 2.5 kilobases and highly correlated (absolute Pearson correlation >0.5) with a lead CpG that had posterior inclusion probability greater than 20% were grouped together. A lead probe was defined as having an initial posterior inclusion probability of greater than 20%. For each probe group, we calculated the proportion of iterations for which at least one probe was included in the model, yielding the group posterior inclusion probability. Included in the model refers to the CpG being assigned to the small, medium or large effect group in a given iteration (i.e. it was not in group 0, corresponding to having no effect on the phenotype). We then calculated the mean (across 1,000 iterations) of the sum of squared regression coefficients for the probe group to give the contribution of the group to the total variance. Finally, we took groups where their combined posterior inclusion probability was >80% (i.e. at least one member was present in 950 iterations). We highlighted the lead CpG in each group for clarity. The variance components estimates were taken as the mean sum of squared standardised mean posterior effect sizes across the 1,000 iterations, and the 2.5%ile and 97.5%ile (iteration rank 25 and 975) formed the bounds of the 95% credible interval.

**Variance component estimation – combined analysis**

Genotyping in Generation Scotland was performed using Illumina HumanOmniExpressExome-8 v1.0 Bead Chip or Illumina HumanOmniExpressExome-8 v1.2 Bead Chip. Single nucleotide polymorphisms (SNPs) were excluded on the basis of missing genotype call rate (>2%), departure from the Hardy–Weinberg equilibrium (*p* < 1 × 10^−6^) and low minor allele frequency (<1%). Duplicate samples were removed alongside individuals with sex mismatches and missing genotype call rates (>2%). Principal component analysis was performed on GS samples that were combined with 1,092 individuals of the 1000 Genomes (1000G) population^1^. Outliers were defined as those individuals who were more than six standard deviations away from the mean component for the first two principal components. Genomic distance outliers were excluded from all analyses.

In BayesR+, additively coded genotypes (i.e. 0, 1 or 2 alleles) at 561,125 SNPs were scaled to mean zero and unit variance as in the strategy described within the main text for methylation. Missing genotypes were mean imputed. The same prior mixture variances were used as in the methylation-based analysis i.e. 0.001, 0.01 and 0.1. Identical phenotype preparations were applied. BayesR+ returned estimates for the proportion of variance in CRP attributed to genome-wide methylation and genetic factors when considered alone and also when conditioned on one another.

In OSCA, a sparse genomic relationship matrix was estimated using genotype data (n=561,125 markers, --grm-cutoff 0.05). A GRM was initially fitted alone in order to obtain a heritability estimate for CRP. The –multi-orm flag was used in order to fit an ORM (methylation) and a GRM (genetics) as random effect components for restricted maximum likelihood estimation. This allowed for a joint estimation of epigenetic and genetic variance components.

**Assessing longitudinal stability of CRP measures**

The LBC1936 cohort contained repeat DNAm and CRP measures over four time-points. This cohort had three high-sensitivity measurements at age 73, 76 and 79 years. The intra-class correlation coefficients of all five DNAm CRP predictors were assessed and compared to that of phenotypic CRP. The *ICC* function in the R package *psych* was used and the ‘average random raters’ (ICC2k) model was selected to estimate the temporal stabilities of the relevant measures^2^.

**Assessing relationship between assay-measured or DNAm CRP and future CRP measurements**

We used the R package *lmerTest* to fit linear mixed-effects models and to regress high-sensitivity CRP measurements on an interaction term between CRP (assay-measured or DNAm) at Wave 2 and chronological age^3^. We co-varied for sex and fitted participant ID as a random effect on the intercept. Here, CRP (assay-measured or DNAm) at Wave 2 was used as the baseline measurement given that Wave 1 contains low-sensitivity measurements alone. Waves 3 and Wave 4 also contain high-sensitivity measurements allowing for a fair comparison of longitudinal relationships across Waves 2, 3 and 4.

**Polygenic score profiling in Lothian Birth Cohort 1936**

A polygenic or genetic score for CRP was computed in LBC1936 participants using PRSice-2 software^4^. LBC1936 DNA samples were genotyped at the Edinburgh Clinical Research Facility using the Illumina 610-Quadv1 array (Wave 1; n = 1,005; mean age: 69.6 ± 0.8 years; San Diego)^5^. SNPs were imputed to the 1000G reference panel (phase 3, version 5)^6^. Individuals were excluded on the basis of sex mismatches, family structure, SNP call rates below 95%, and evidence of non-European ancestry. SNPs with a call rate of greater than 98%, minor allele frequency in excess of 1%, and Hardy-Weinberg equilibrium test with *p* ≥ 0.001 were retained. An imputation quality score of > 0.8 was applied to the imputed set of variants.

Summary statistics from a recent, large genome-wide association study on CRP levels were applied to build the genetic risk score^7^. Only summary data from European samples were applied. An additive weighted genetic score for CRP was constructed from SNPs that passed the genome-wide threshold in the association study (*p* < 5 × 10^-8^). Weighted dosages were calculated by multiplying the dose of each risk allele by the effect estimate from the GWAS. The sum of these products produced a genetic risk score for each individual.

**Note on elastic net regression**

It is an intermediary of other penalised regression methods, which may model all features simultaneously to produce parsimonious solutions that account for probe correlations (e.g., LASSO regression) or those that apply small weights to all features (e.g., ridge regression).
