## Additional file 4 for "Blood-based epigenome-wide analyses of chronic low-grade inflammation across diverse population cohorts"

**Additional file 4 - Supplementary Figures.** The following information pertains to Supplementary Figures for the manuscript ‘*Blood-based epigenome-wide analyses of chronic low-grade inflammation across diverse population cohorts’* by Hillary *et al.*

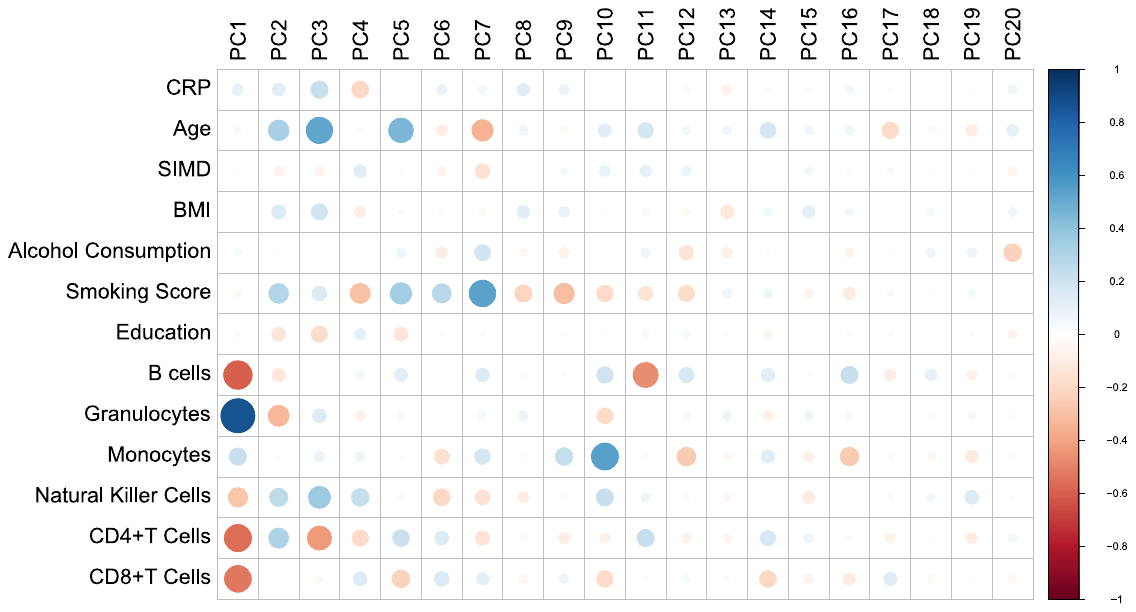

**Fig. S1. Correlation between the first 20 principal components on CRP-associated probes and continuous covariates in Generation Scotland.** Principal component analysis was applied to a set of 1,379 CRP-associated probes from Wielscher *et al.* in Generation Scotland (N=17,936). Only associations for the first twenty principal components are shown for clarity. BMI, body mass index; CRP, C-reactive protein; PC, principal component; SIMD, Scottish Index of Multiple Deprivation.

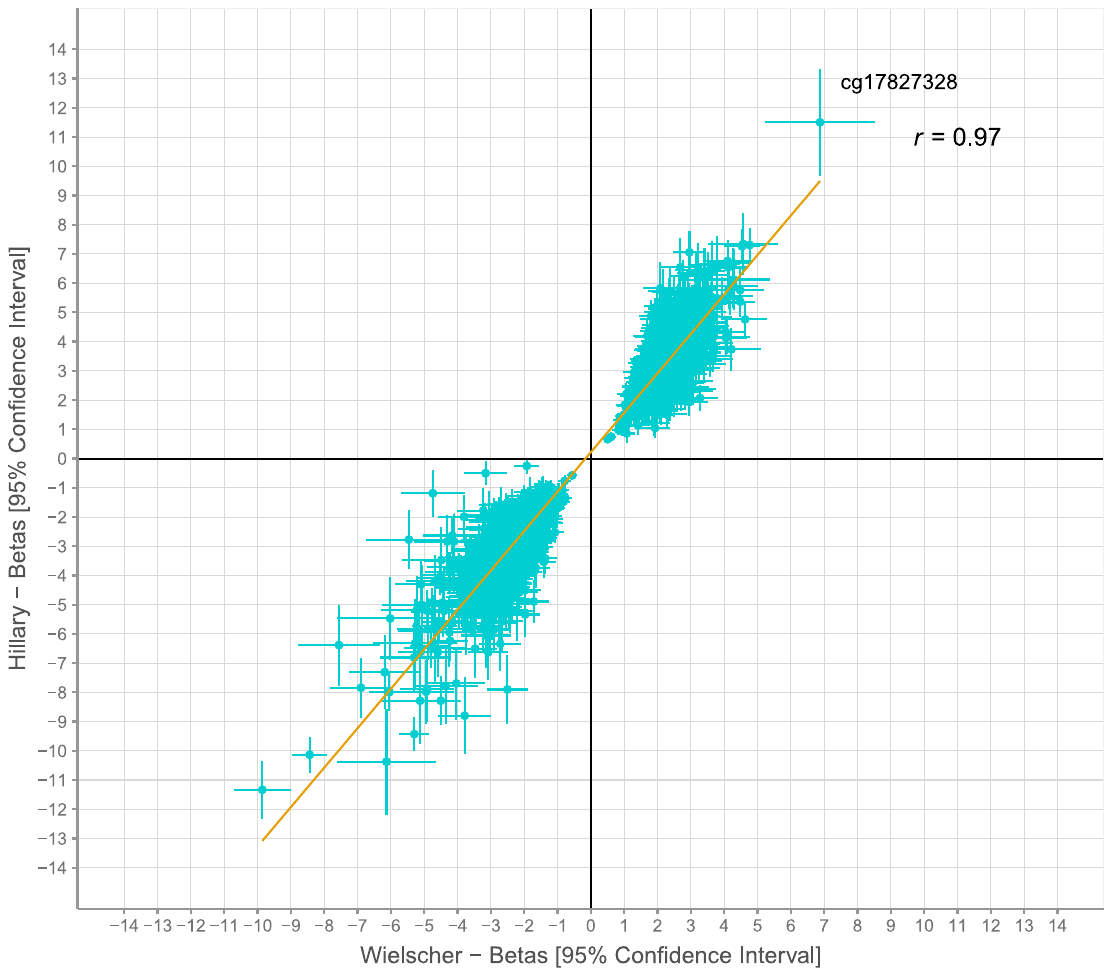

**Fig. S2. Correlation of effect sizes between epigenome-wide association studies by Wielscher *et al.* and the present study (Hillary *et al.*).** Outputs from a basic model are shown, with matched analytical strategies between the two studies. Effect sizes for CpG sites that were significantly associated with blood CRP levels in the Wielscher *et al.* study (at *p* < 3.6 x10^-8^) were compared against corresponding effects in the present study. There were 1,379 such CpG sites common to both studies that underpinned the correlation test. CpG, cytosine-phosphate-guanine dinucleotide; CRP, C-reactive protein.

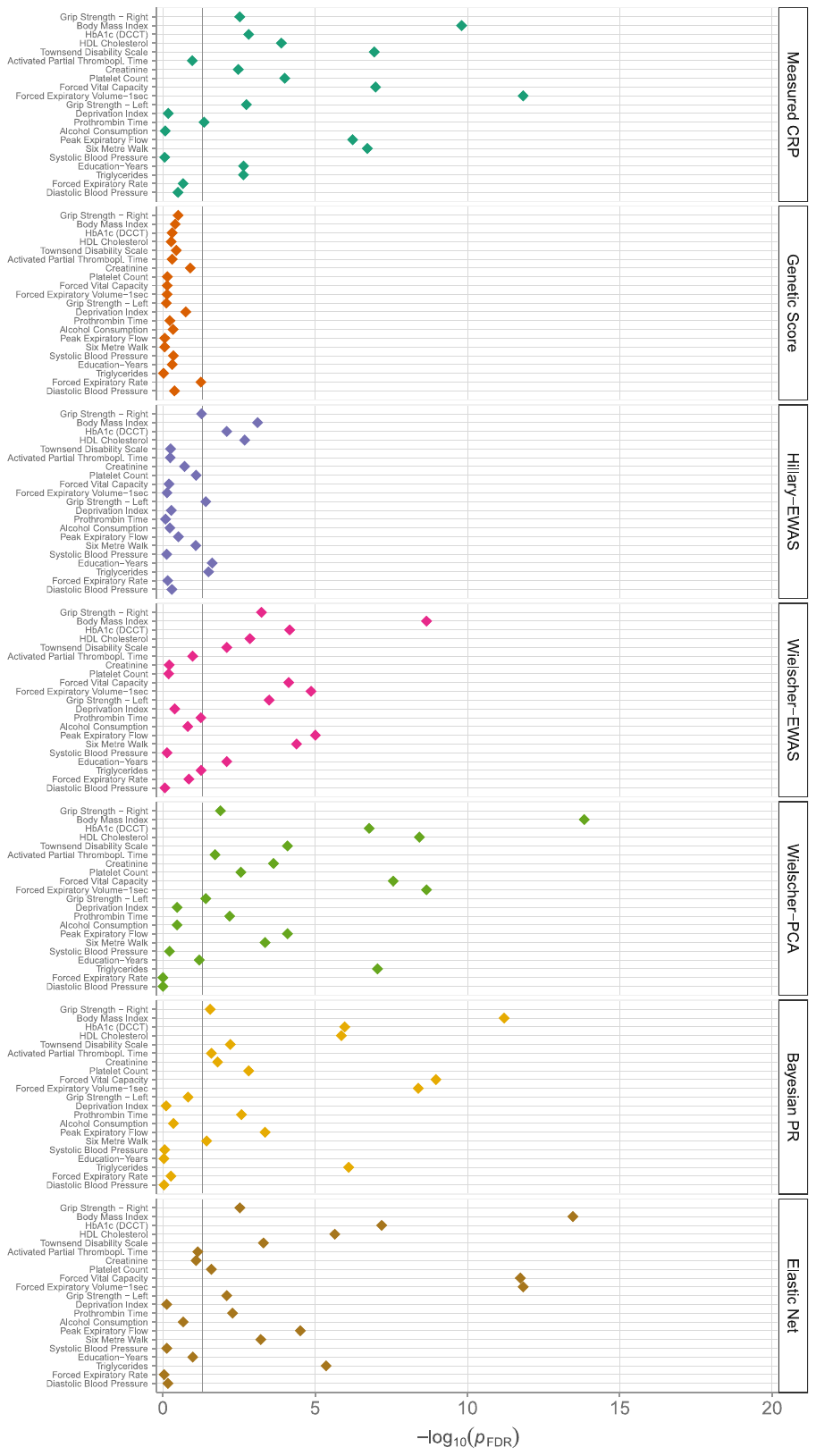

**Fig. S3. Association of 21 continuous cardiometabolic and lifestyle variables with measured CRP, genetic score for CRP and five DNAm predictors of CRP.** CRP, C-reactive protein; DCCT; Diabetes Control and Complications Trial; DNAm, DNA methylation; EWAS, epigenome-wide association study; FDR, false discovery rate; HDL, high-density lipoprotein; PCA, principal component analysis; PR, penalised regression; Thrombopl., thromboplastin.

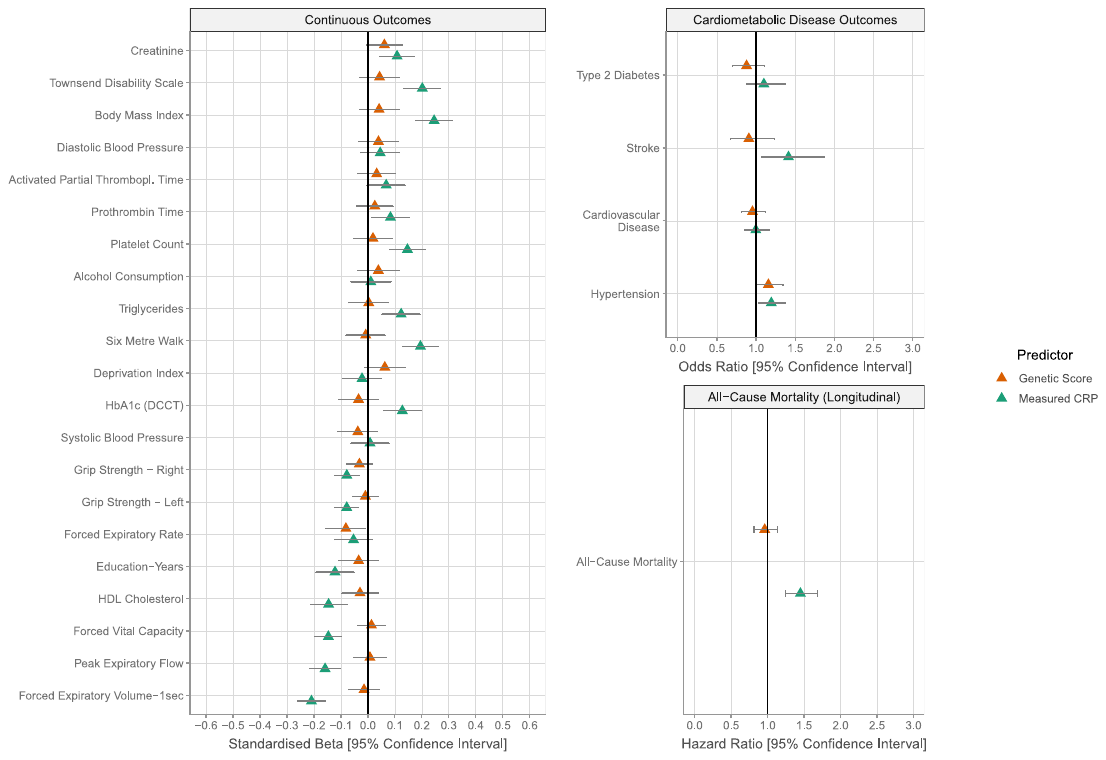

**Fig. S4. Associations of health outcomes with a genetic score for CRP or assay-measured CRP in the Lothian Birth Cohort 1936.** CRP; C-reactive protein; DCCT; Diabetes Control and Complications Trial; DNAm, DNA methylation; HDL, high-density lipoprotein; Thrombopl., thromboplastin.

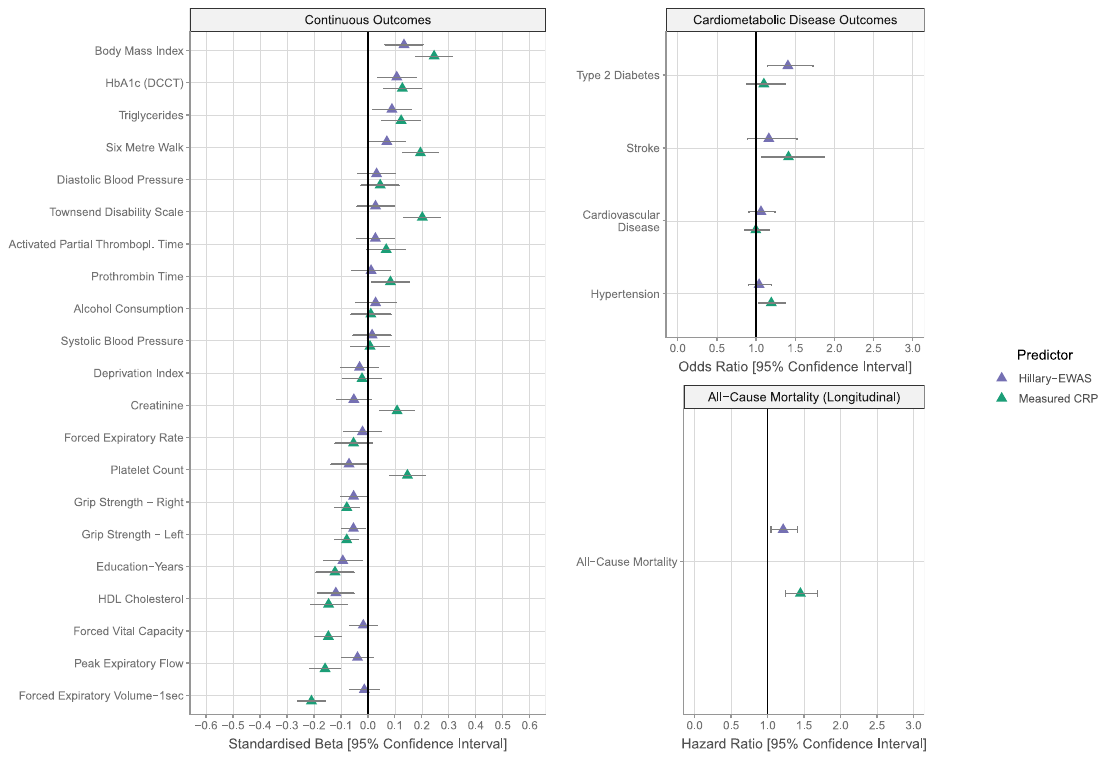

**Fig. S5. Associations of health outcomes with DNAm CRP (Hillary EWAS-based predictor) or assay-measured CRP in the Lothian Birth Cohort 1936.** CRP; C-reactive protein; DCCT; Diabetes Control and Complications Trial; DNAm, DNA methylation; EWAS, epigenome-wide association study; HDL, high-density lipoprotein; Thrombopl., thromboplastin.

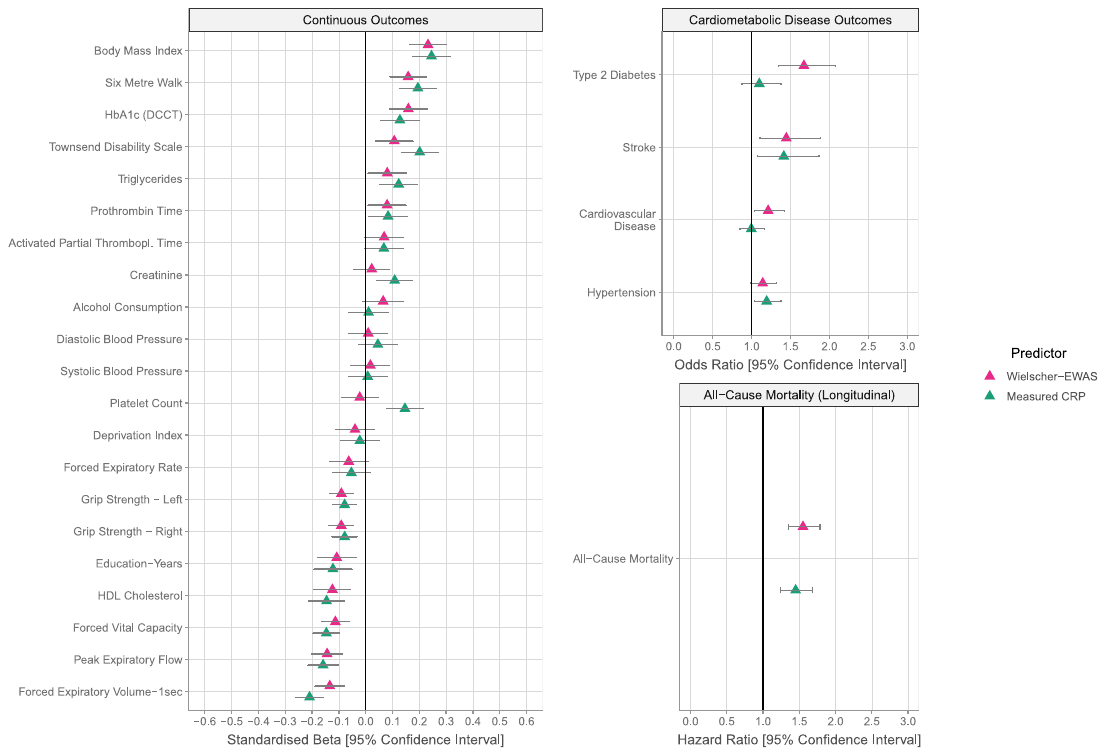

**Fig. S6. Associations of health outcomes with DNAm CRP (Wielscher EWAS-based predictor) or assay-measured CRP in the Lothian Birth Cohort 1936.** CRP; C-reactive protein; DCCT; Diabetes Control and Complications Trial; DNAm, DNA methylation; EWAS, epigenome-wide association study; HDL, high-density lipoprotein; Thrombopl., thromboplastin.

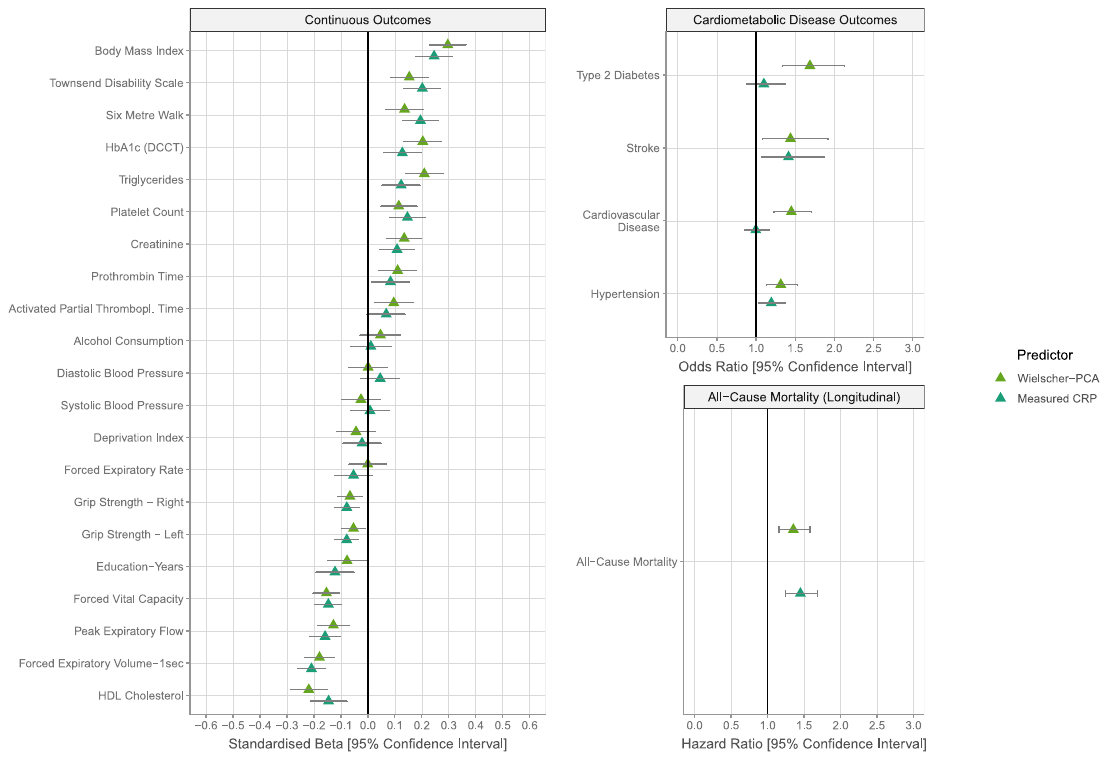

**Fig. S7. Associations of health outcomes with DNAm CRP (PCA-based predictor) or assay-measured CRP in the Lothian Birth Cohort 1936.** CRP; C-reactive protein; DCCT; Diabetes Control and Complications Trial; DNAm, DNA methylation; HDL, high-density lipoprotein; PCA, principal component analysis; Thrombopl., thromboplastin.

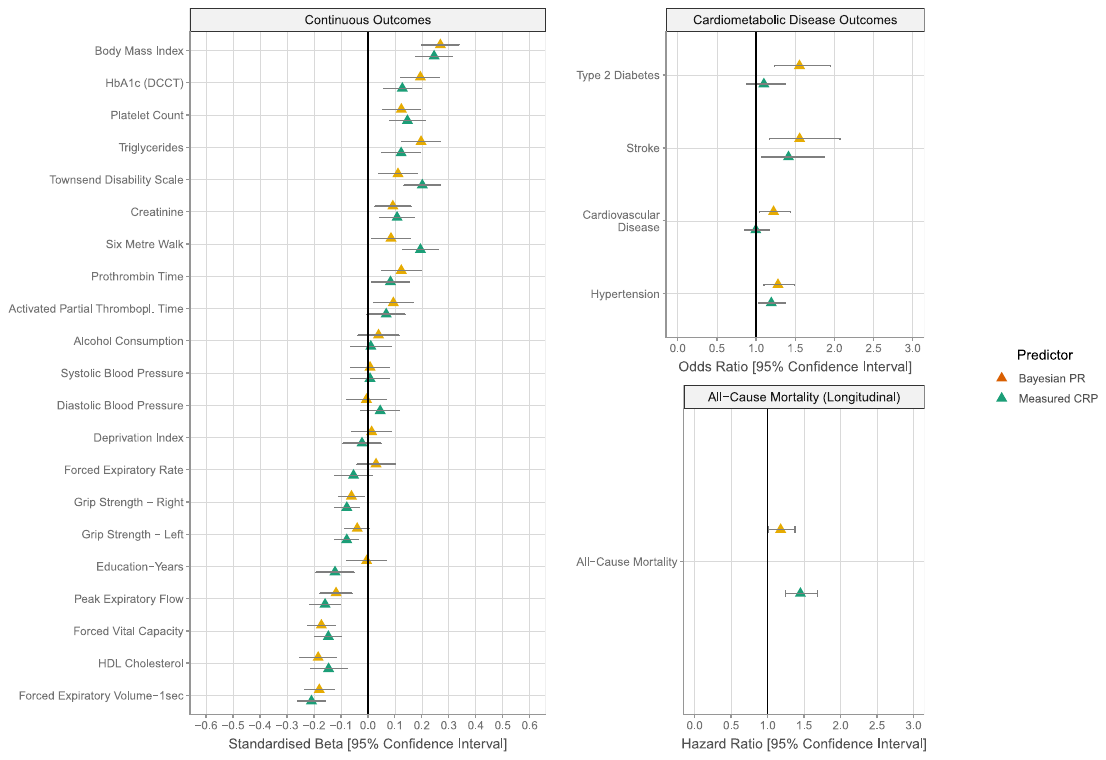

**Fig. S8. Associations of health outcomes with DNAm CRP (Bayesian PR-based predictor) or assay-measured CRP in the Lothian Birth Cohort 1936.** CRP; C-reactive protein; DCCT; Diabetes Control and Complications Trial; DNAm, DNA methylation; HDL, high-density lipoprotein; PR, penalised regression; Thrombopl., thromboplastin.
