## Additional file 5 for "Blood-based epigenome-wide analyses of chronic low-grade inflammation across diverse population cohorts"

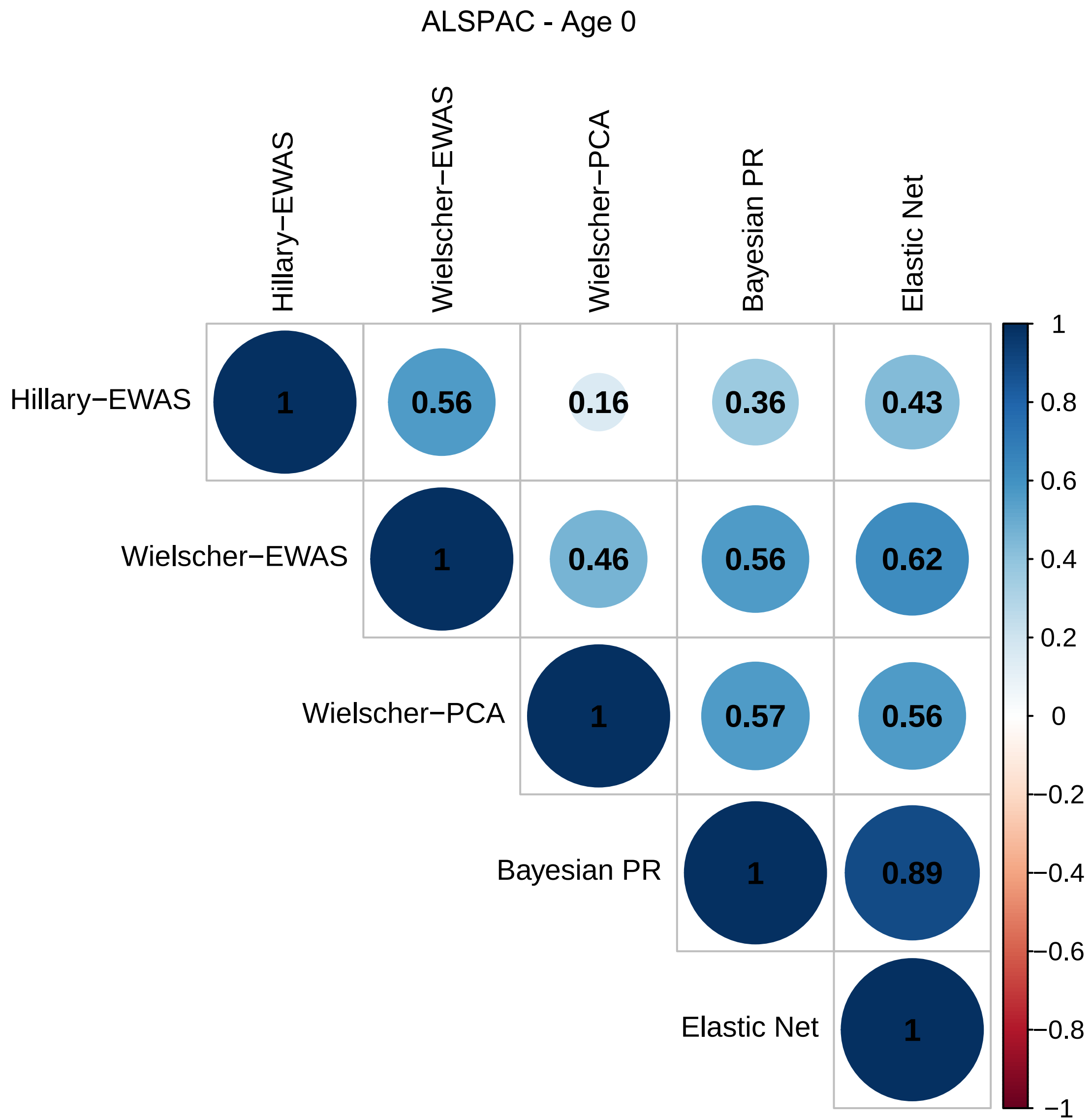

ALSPAC - Age 0
Hillary−EWAS
Wielscher−EWAS
Wielscher−PCA
Bayesian PR
Elastic Net
1
0.56
0.16
0.36
0.43
1
0.46
0.56
0.62
1
0.57
0.56
1
0.89
1
1
0.8
0.6
0.4
0.2
0
−0.2
−0.4
−0.6
−0.8
−1

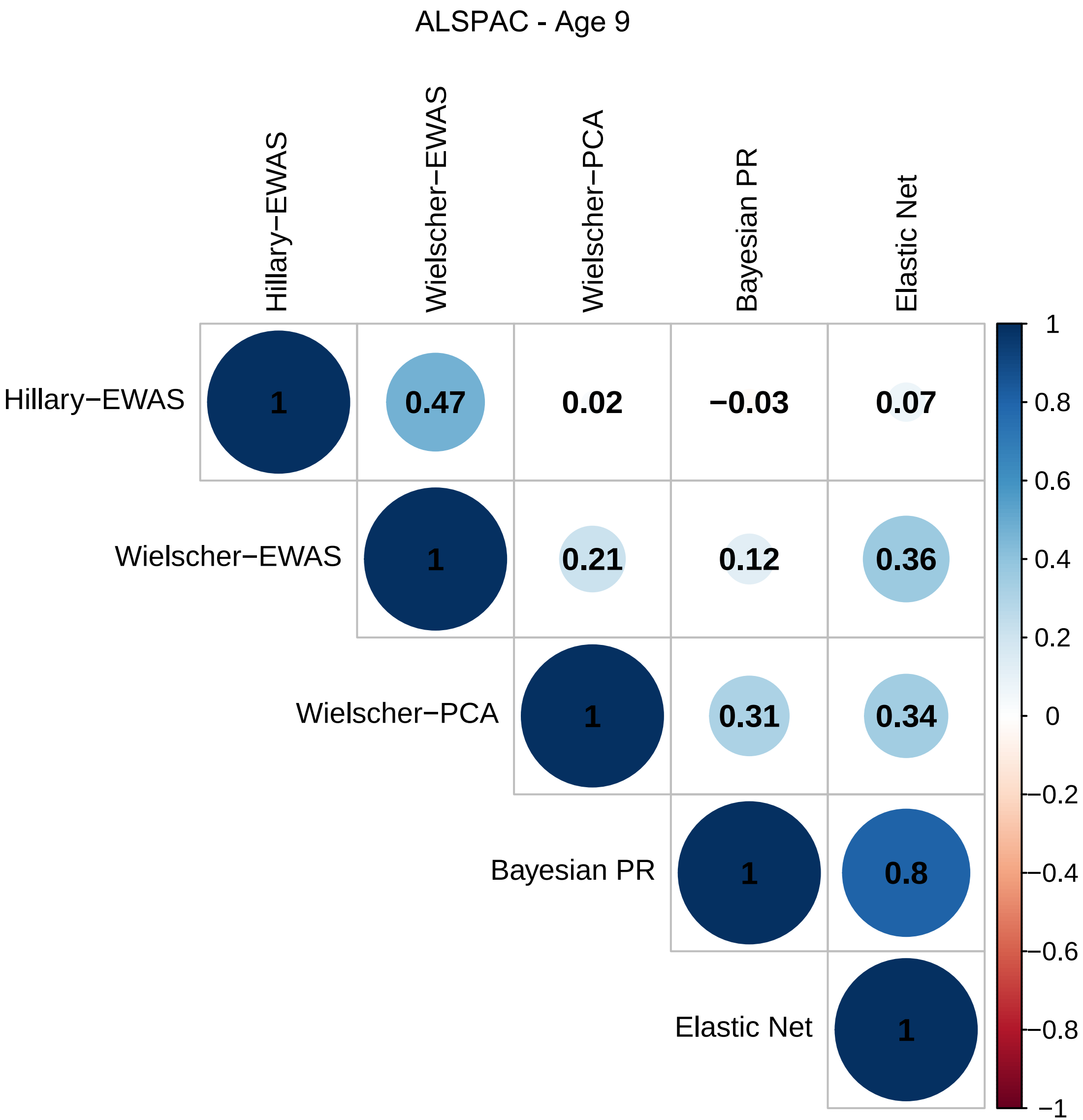
ALSPAC - Age 9
Hillary−EWAS
Wielscher−EWAS
Wielscher−PCA
Bayesian PR
Elastic Net
1
0.47
0.02
−0.03
0.07
1
0.21
0.12
0.36
1
0.31
0.34
1
0.8
1
1
0.8
0.6
0.4
0.2
0
−0.2
−0.4
−0.6
−0.8
−1

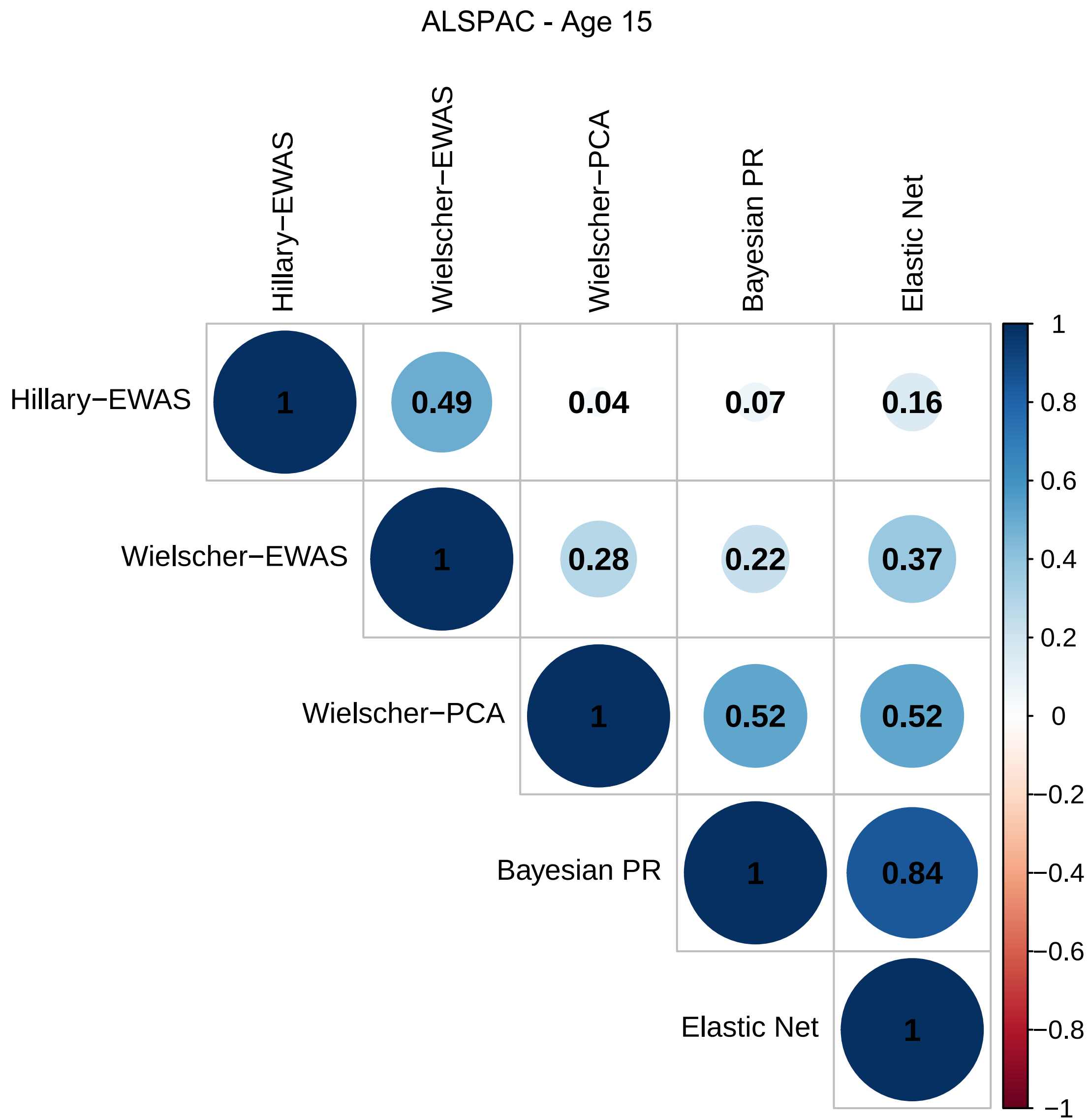
ALSPAC - Age 15
Hillary−EWAS
Wielscher−EWAS
Wielscher−PCA
Bayesian PR
Elastic Net
1
0.49
0.04
0.07
0.16
1
0.28
0.22
0.37
1
0.52
0.52
1
0.84
1
1
0.8
0.6
0.4
0.2
0
−0.2
−0.4
−0.6
−0.8
−1

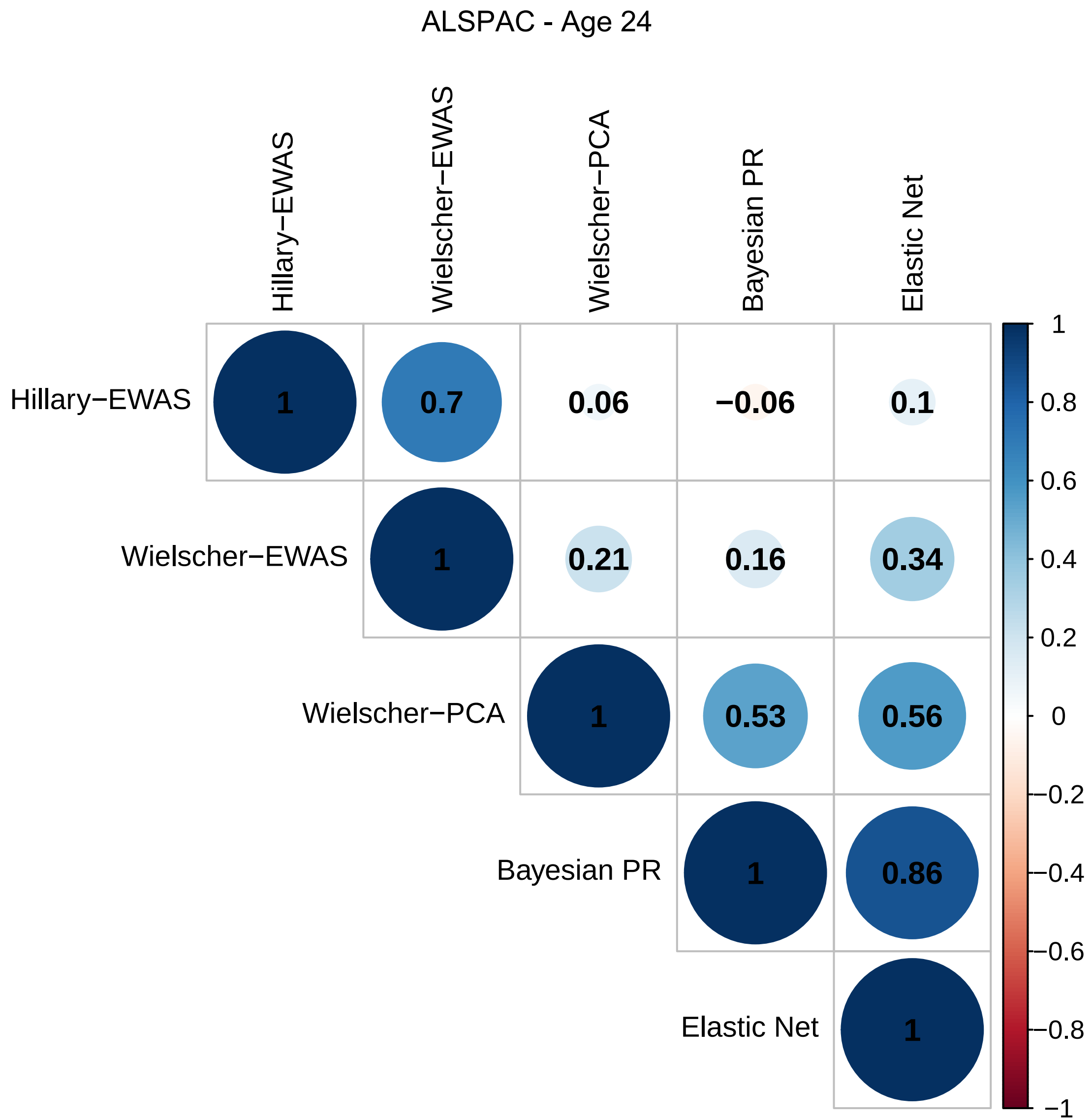
ALSPAC - Age 24
Hillary−EWAS
Wielscher−EWAS
Wielscher−PCA
Bayesian PR
Elastic Net
1
0.7
0.06
−0.06
0.1
1
0.21
0.16
0.34
1
0.53
0.56
1
0.86
1
1
0.8
0.6
0.4
0.2
0
−0.2
−0.4
−0.6
−0.8
−1

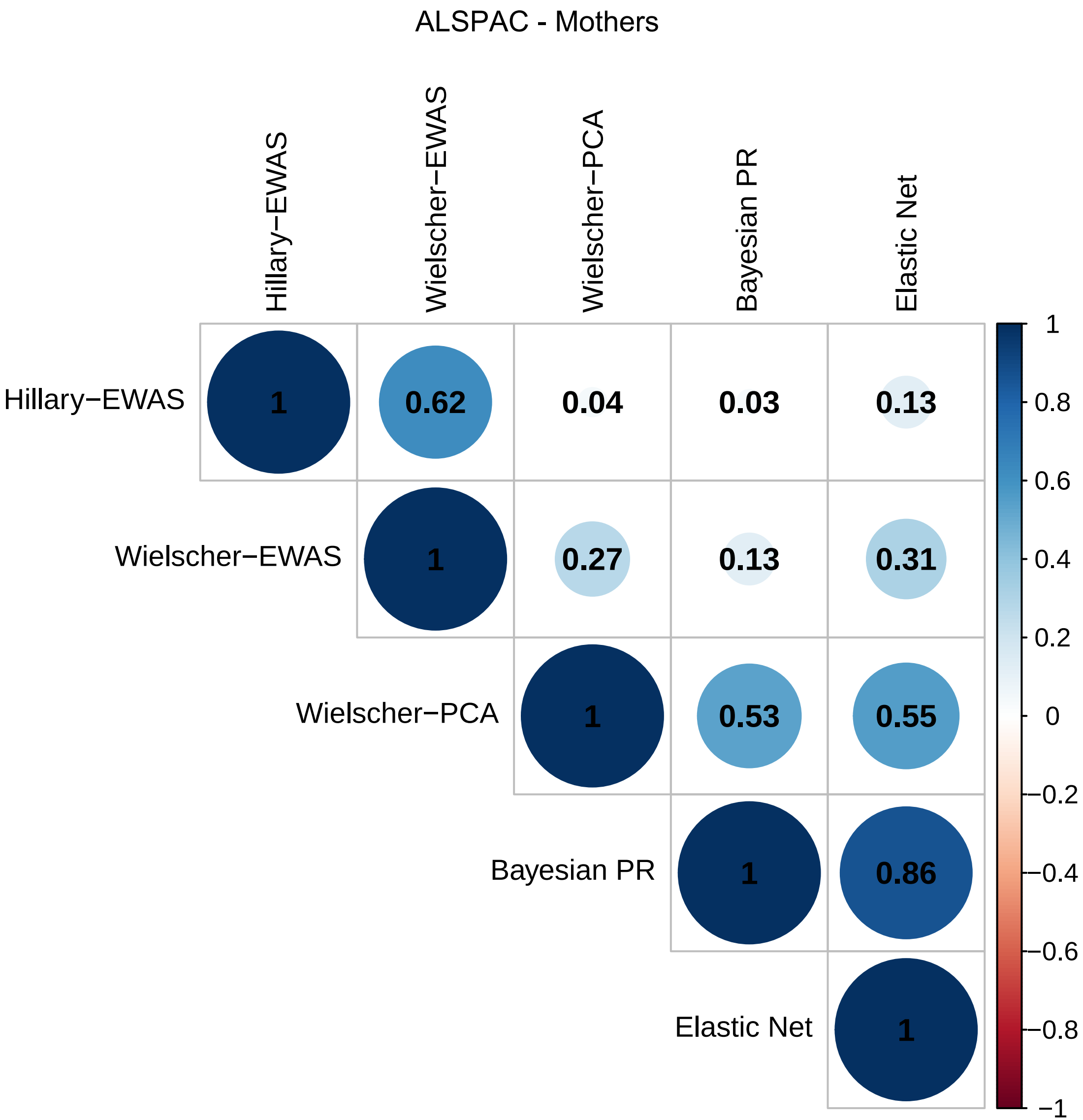

ALSPAC - Mothers
Hillary−EWAS
Wielscher−EWAS
Wielscher−PCA
Bayesian PR
Elastic Net
1
0.62
0.04
0.03
0.13
1
0.27
0.13
0.31
1
0.53
0.55
1
0.86
1
1
0.8
0.6
0.4
0.2
0
−0.2
−0.4
−0.6
−0.8
−1

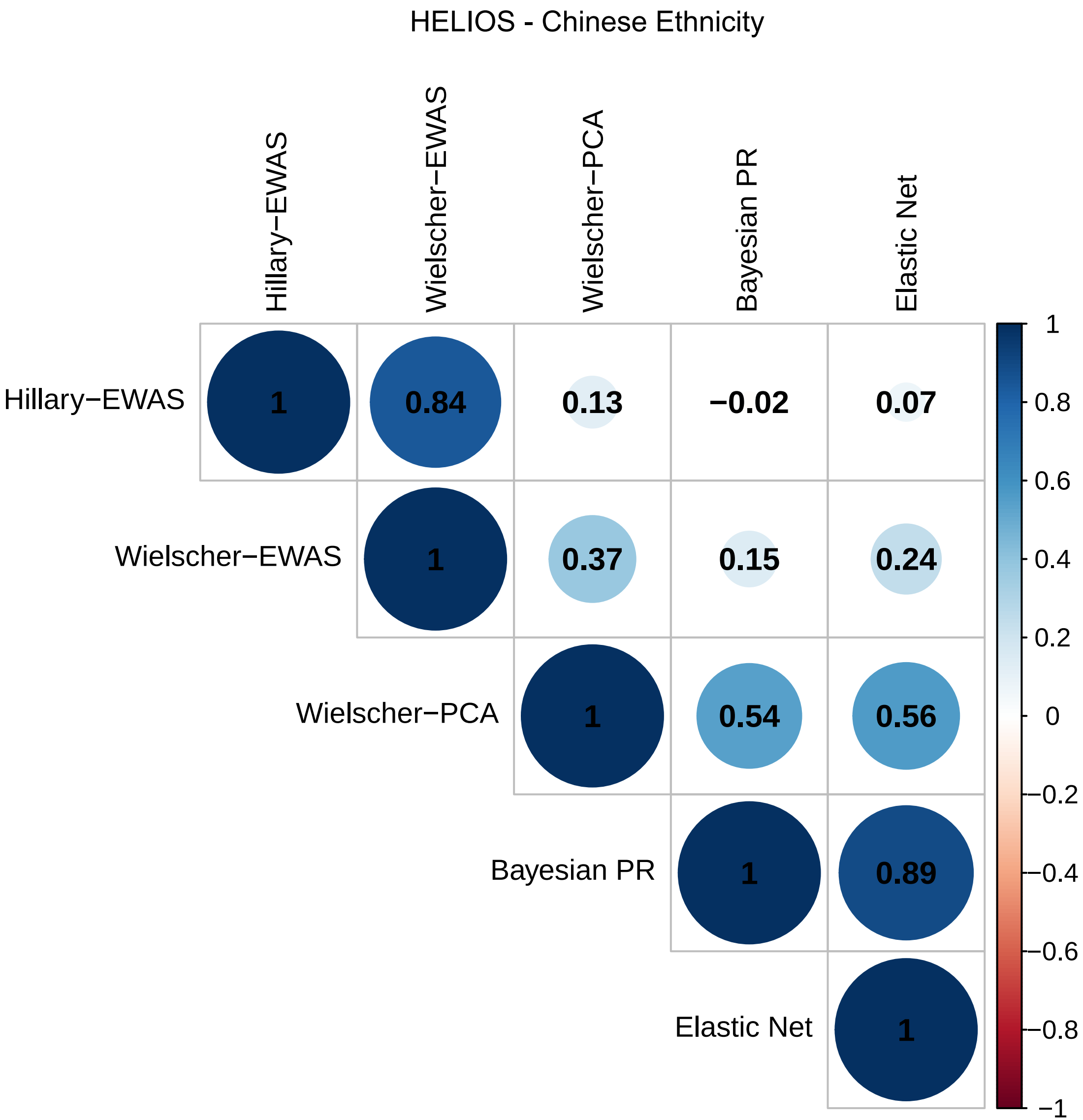
HELIOS - Chinese Ethnicity
Hillary−EWAS
Wielscher−EWAS
Wielscher−PCA
Bayesian PR
Elastic Net
1
0.84
0.13
−0.02
0.07
1
0.37
0.15
0.24
1
0.54
0.56
1
0.89
1
1
0.8
0.6
0.4
0.2
0
−0.2
−0.4
−0.6
−0.8
−1

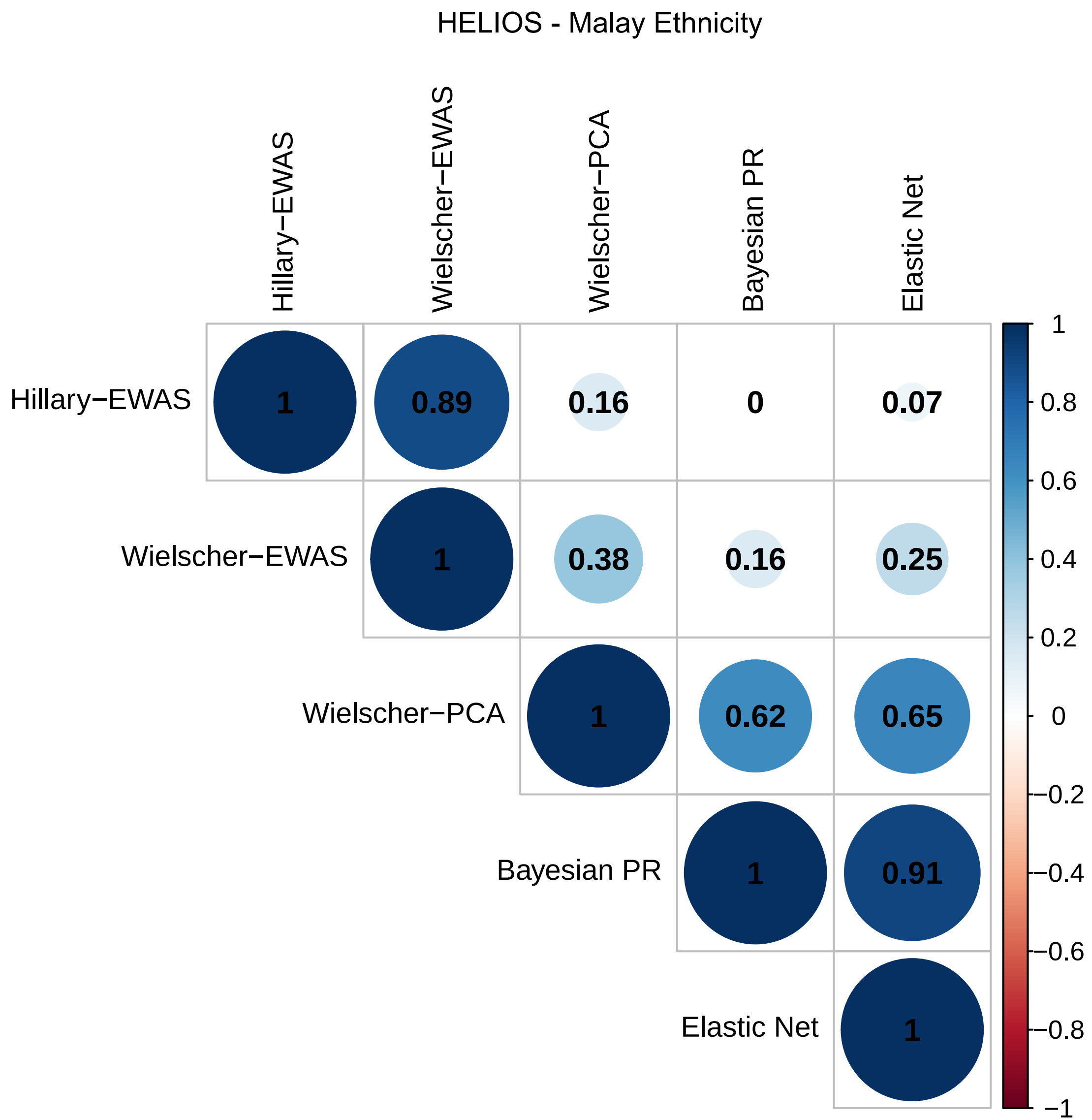
HELIOS - Malay Ethnicity
Hillary−EWAS
Wielscher−EWAS
Wielscher−PCA
Bayesian PR
Elastic Net
1
0.89
0.16
0
0.07
1
0.38
0.16
0.25
1
0.62
0.65
1
0.91
1
1
0.8
0.6
0.4
0.2
0
−0.2
−0.4
−0.6
−0.8
−1

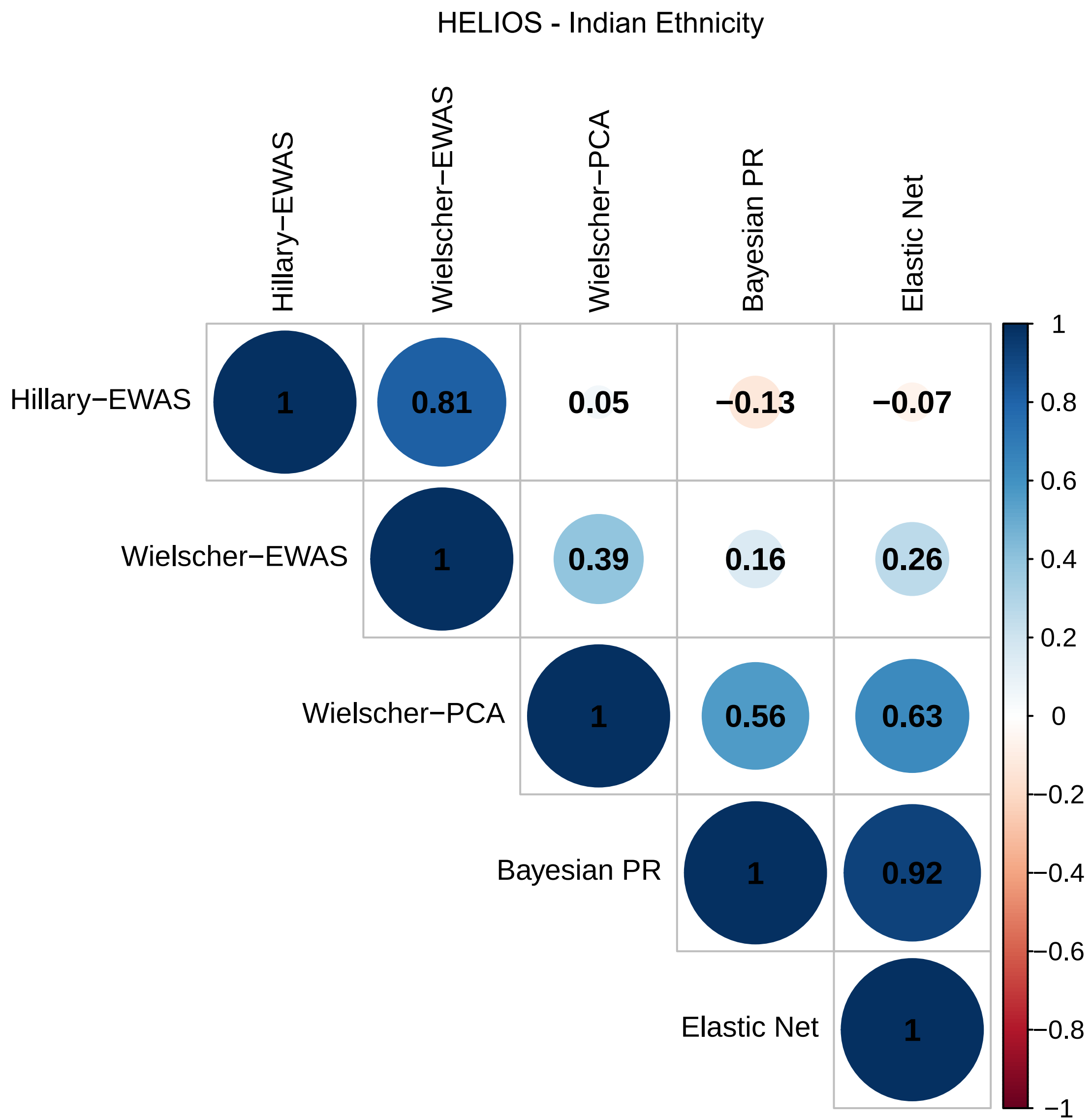
HELIOS - Indian Ethnicity
Hillary−EWAS
Wielscher−EWAS
Wielscher−PCA
Bayesian PR
Elastic Net
1
0.81
0.05
−0.13
−0.07
1
0.39
0.16
0.26
1
0.56
0.63
1
0.92
1
1
0.8
0.6
0.4
0.2
0
−0.2
−0.4
−0.6
−0.8
−1

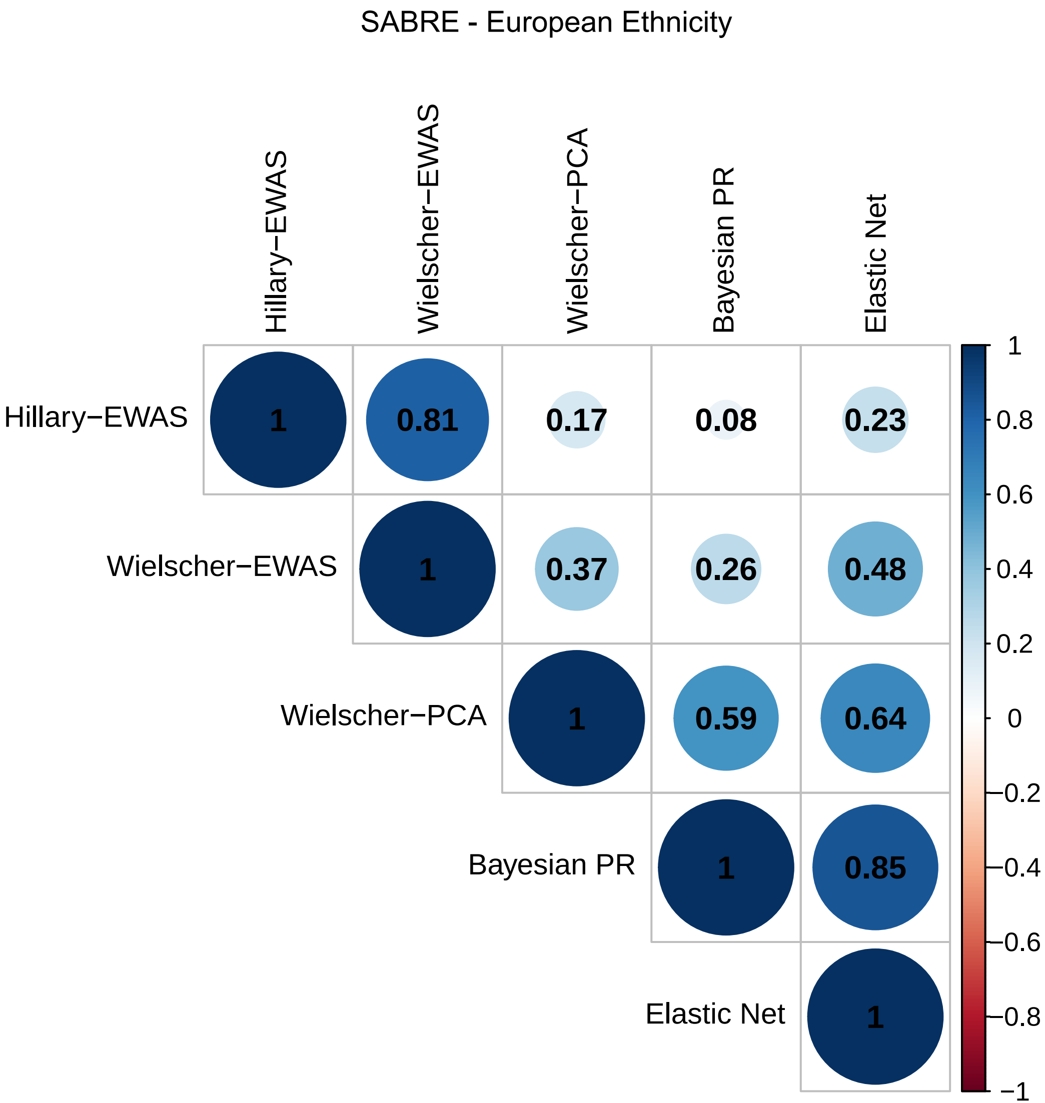

SABRE - European Ethnicity
Hillary−EWAS
Wielscher−EWAS
Wielscher−PCA
Bayesian PR
Elastic Net
1
0.81
0.17
0.08
0.23
1
0.37
0.26
0.48
1
0.59
0.64
1
0.85
1
1
0.8
0.6
0.4
0.2
0
−0.2
−0.4
−0.6
−0.8
−1

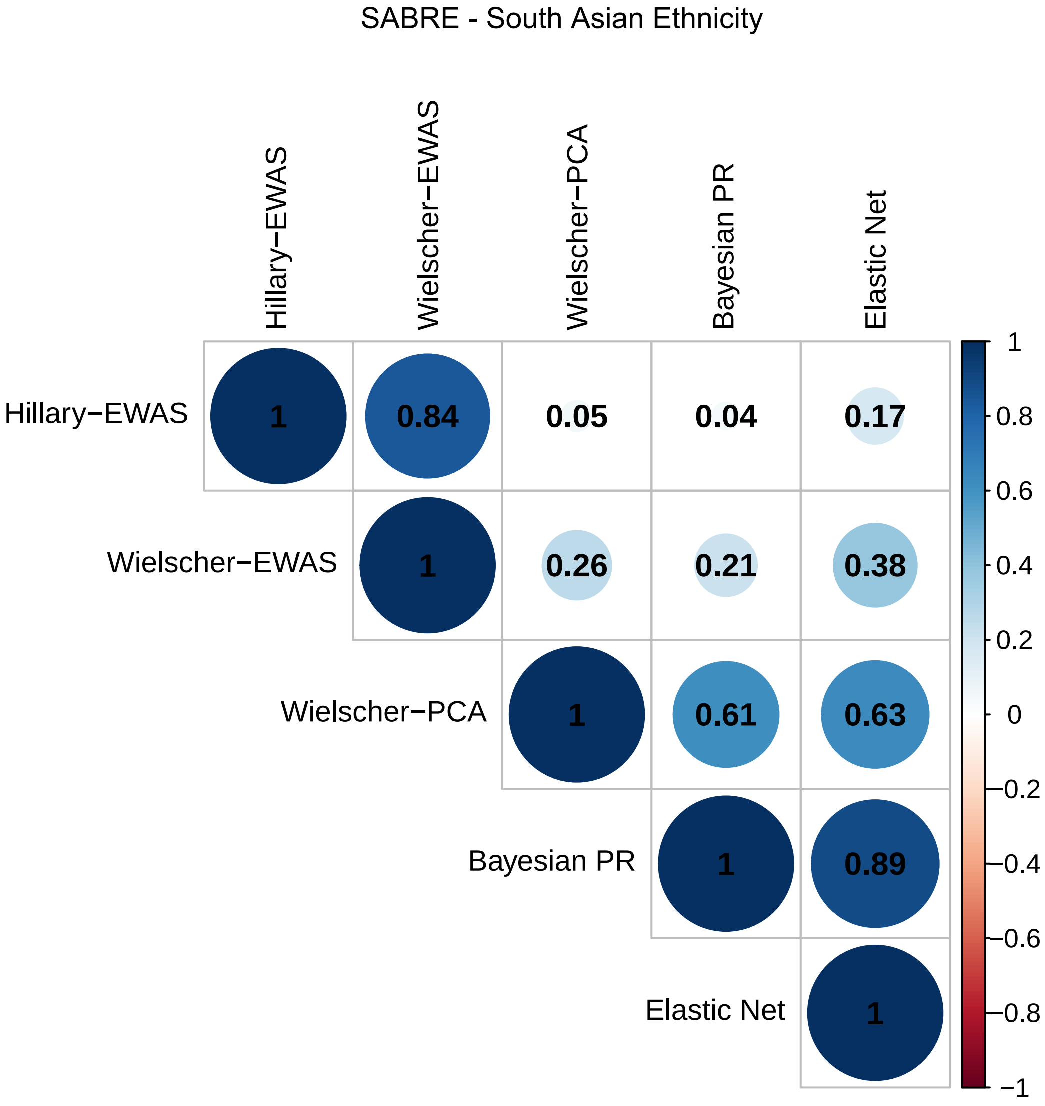
SABRE - South Asian Ethnicity
Hillary−EWAS
Wielscher−EWAS
Wielscher−PCA
Bayesian PR
Elastic Net
1
0.84
0.05
0.04
0.17
1
0.26
0.21
0.38
1
0.61
0.63
1
0.89
1
1
0.8
0.6
0.4
0.2
0
−0.2
−0.4
−0.6
−0.8
−1

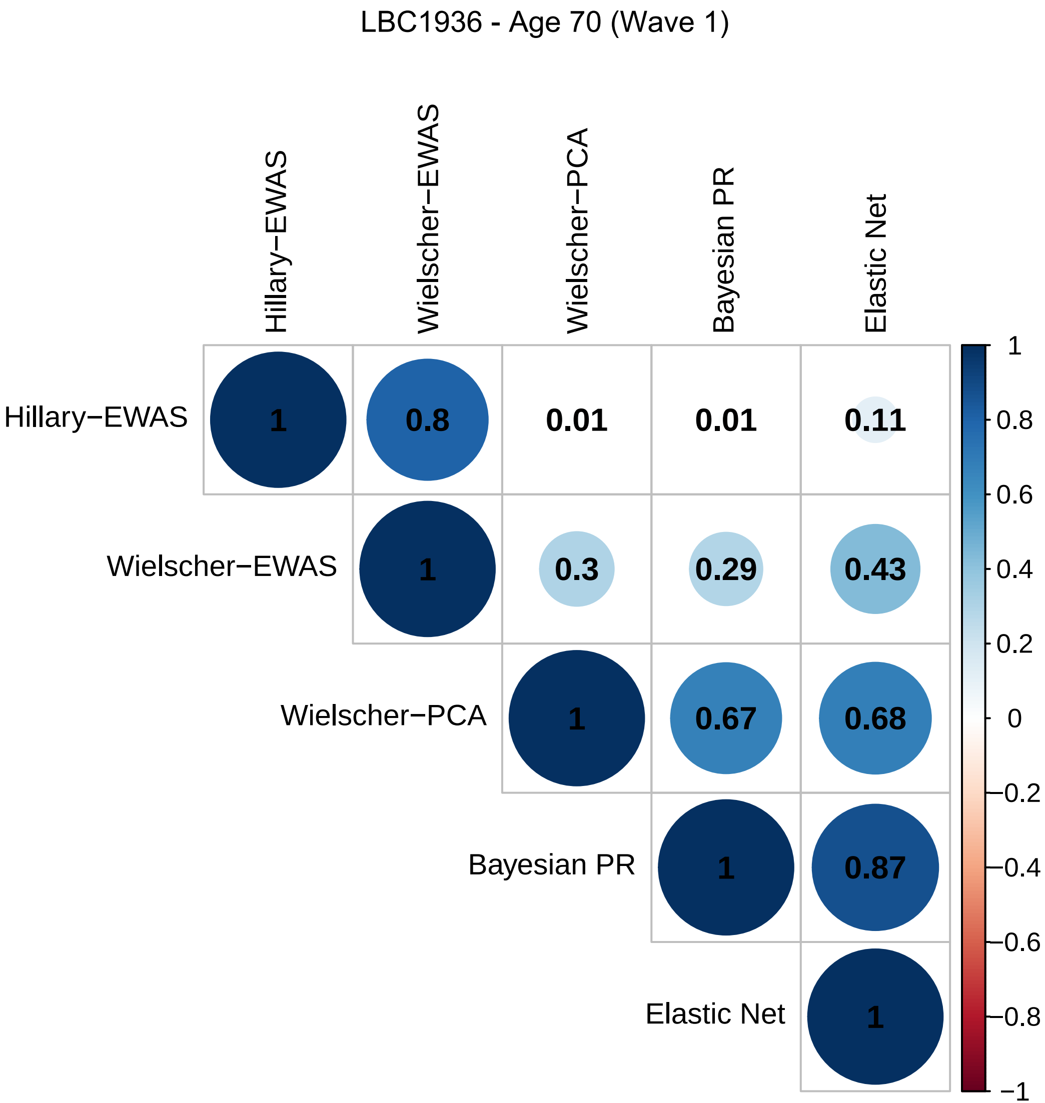
LBC1936 - Age 70 (Wave 1)
Hillary−EWAS
Wielscher−EWAS
Wielscher−PCA
Bayesian PR
Elastic Net
1
0.8
0.01
0.01
0.11
1
0.3
0.29
0.43
1
0.67
0.68
1
0.87
1
1
0.8
0.6
0.4
0.2
0
−0.2
−0.4
−0.6
−0.8
−1

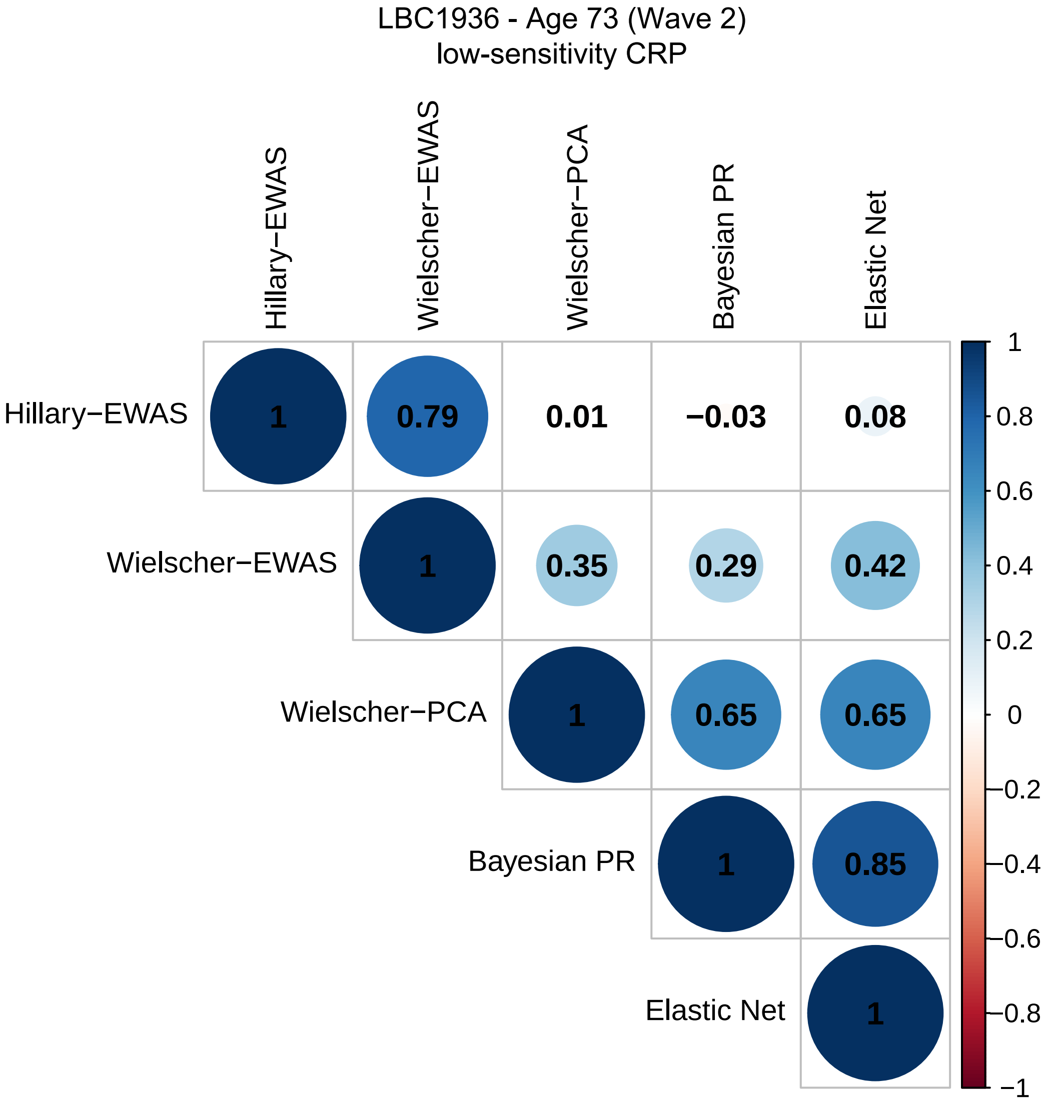

LBC1936 - Age 73 (Wave 2)
low-sensitivity CRP
Hillary−EWAS
Wielscher−EWAS
Wielscher−PCA
Bayesian PR
Elastic Net
1
0.79
0.01
−0.03
0.08
0.35
0.29
0.42
0.65
0.65
0.85
1
0.8
0.6
0.4
0.2
0
−0.2
−0.4
−0.6
−0.8
−1

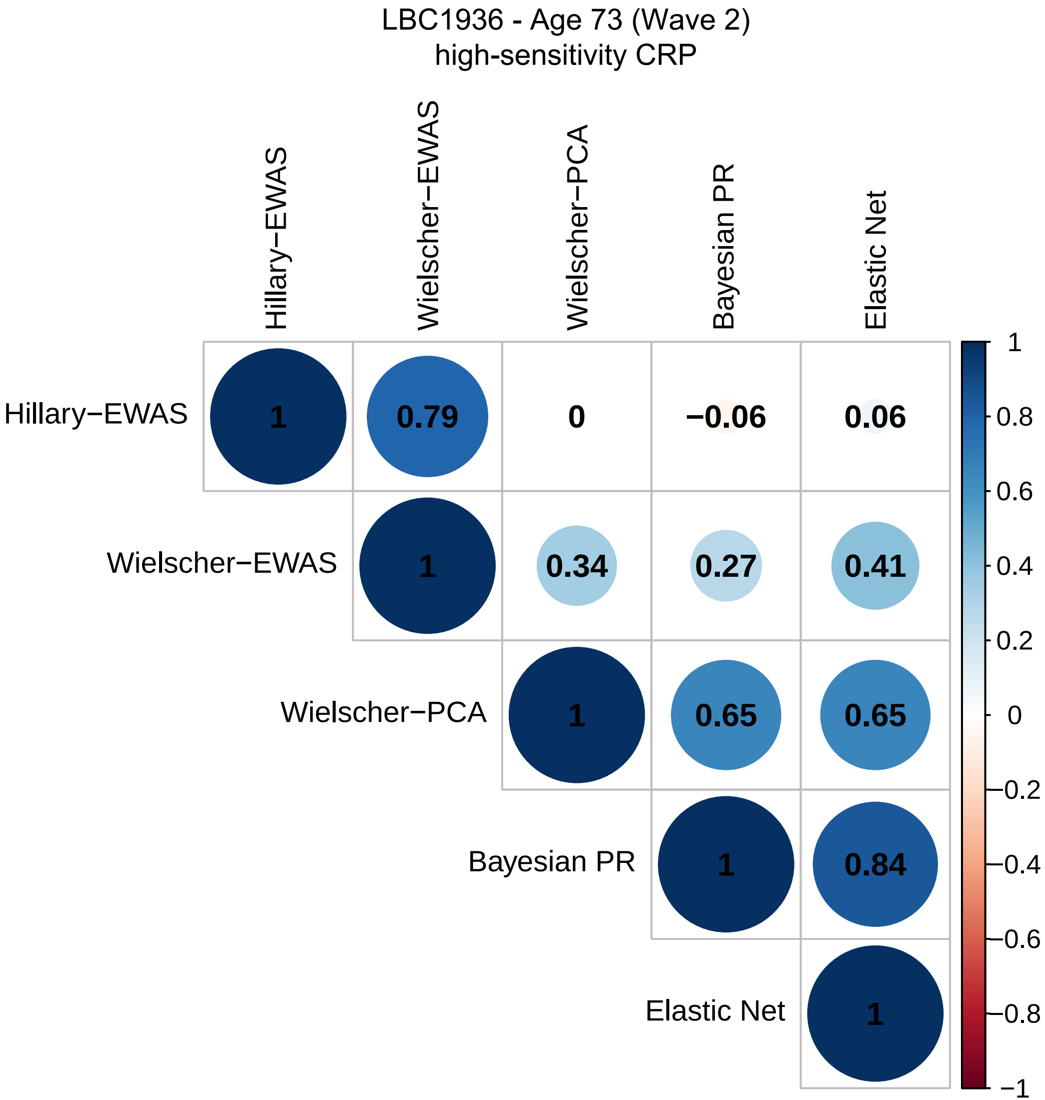
LBC1936 - Age 73 (Wave 2)
high-sensitivity CRP
Hillary−EWAS
Wielscher−EWAS
Wielscher−PCA
Bayesian PR
Elastic Net
1
0.79
0
−0.06
0.06
1
0.34
0.27
0.41
1
0.65
0.65
1
0.84
1
1
0.8
0.6
0.4
0.2
0
−0.2
−0.4
−0.6
−0.8
−1

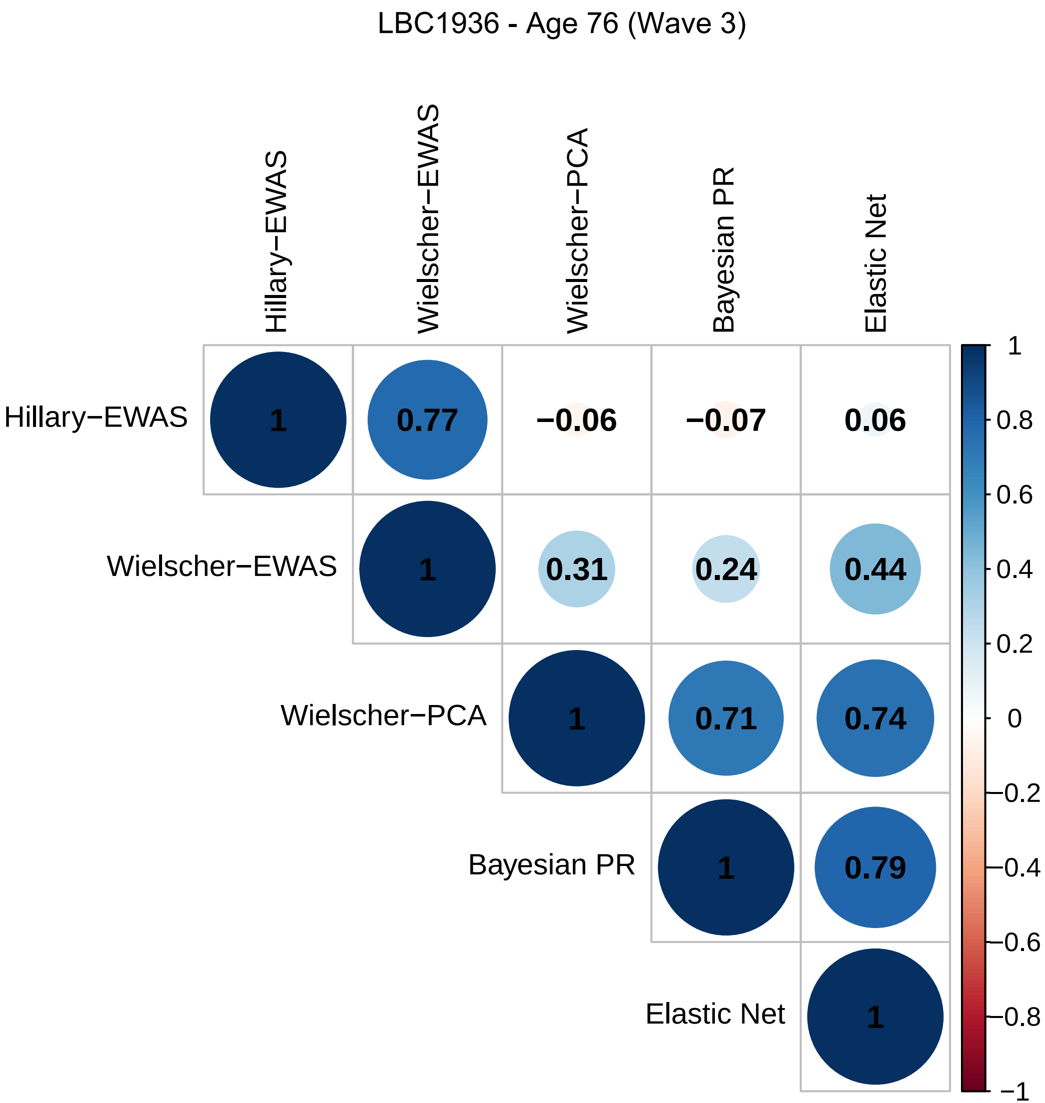
LBC1936 - Age 76 (Wave 3)
Hillary−EWAS
Wielscher−EWAS
Wielscher−PCA
Bayesian PR
Elastic Net
1
0.77
−0.06
−0.07
0.06
1
0.31
0.24
0.44
1
0.71
0.74
1
0.79
1
1
0.8
0.6
0.4
0.2
0
−0.2
−0.4
−0.6
−0.8
−1

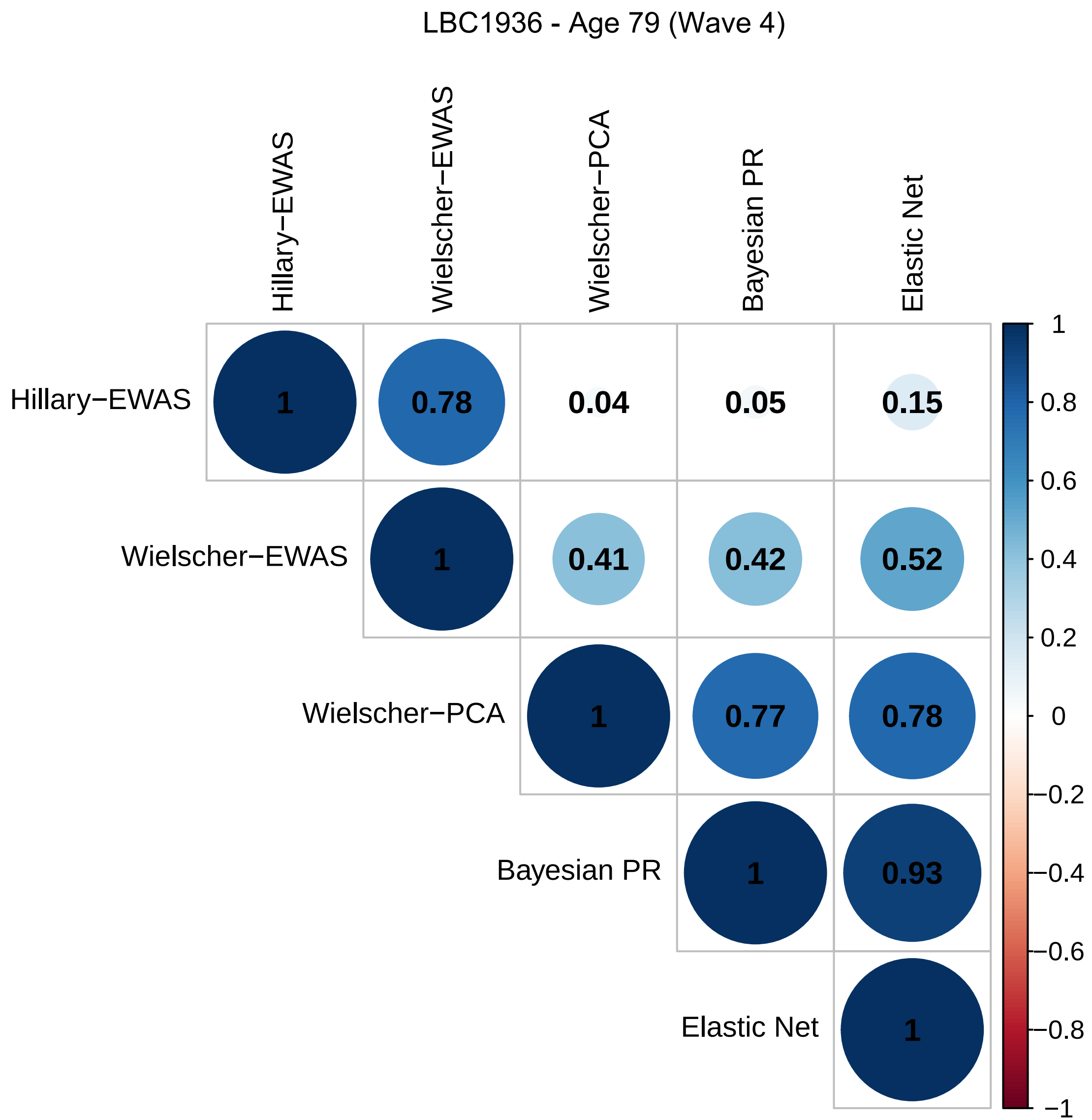
LBC1936 - Age 79 (Wave 4)
Hillary−EWAS
Wielscher−EWAS
Wielscher−PCA
Bayesian PR
Elastic Net
1
0.78
0.04
0.05
0.15
1
0.41
0.42
0.52
1
0.77
0.78
1
0.93
1
1
0.8
0.6
0.4
0.2
0
−0.2
−0.4
−0.6
−0.8
−1

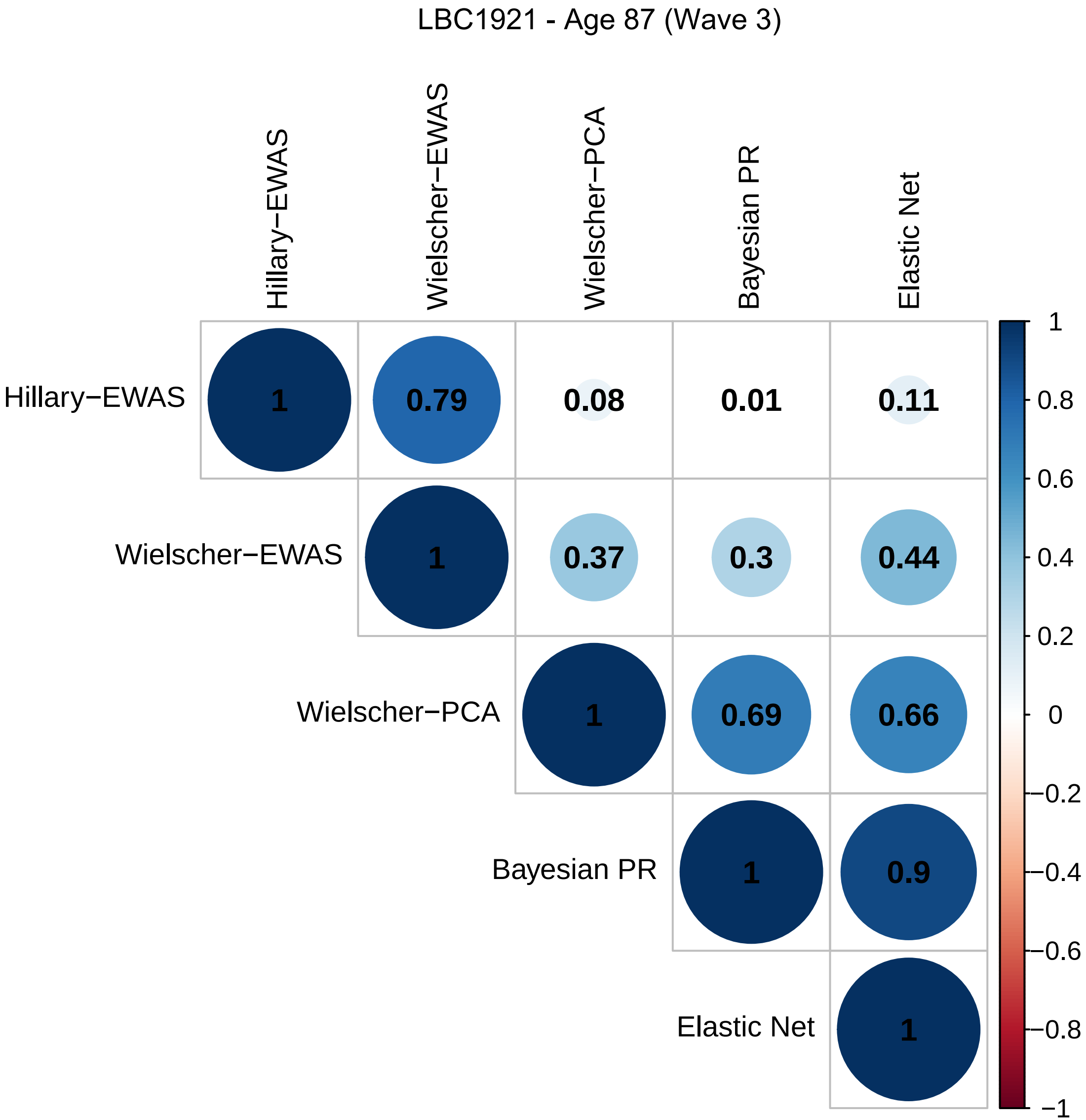
LBC1921 - Age 87 (Wave 3)
Hillary−EWAS
Wielscher−EWAS
Wielscher−PCA
Bayesian PR
Elastic Net
1
0.79
0.08
0.01
0.11
0.37
0.3
0.44
0.69
0.66
0.9
1
0.8
0.6
0.4
0.2
0
−0.2
−0.4
−0.6
−0.8
−1

LBC1921 - Age 90 (Wave 4)

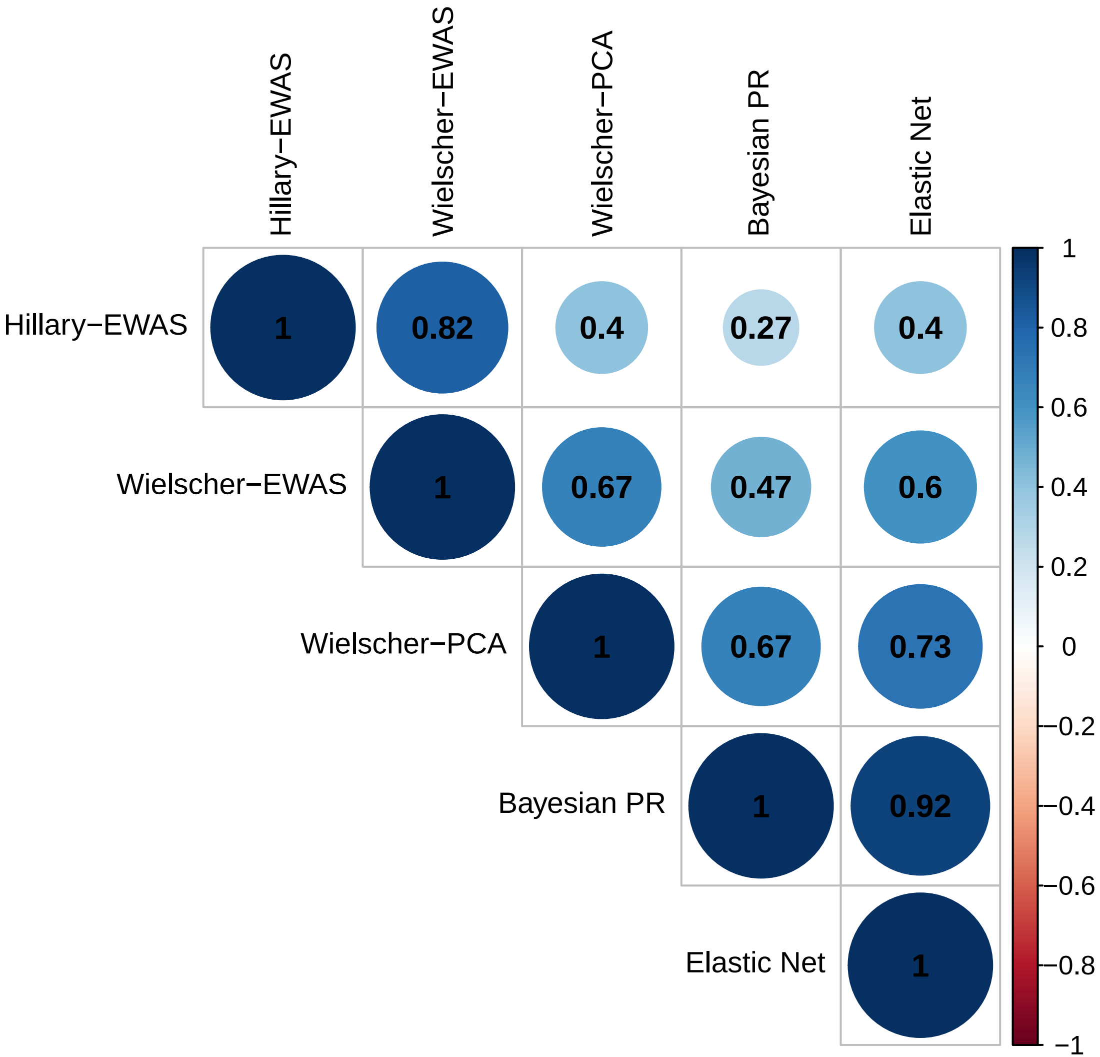
